# A fructo-oligosaccharide prebiotic is well-tolerated in adults undergoing allogeneic hematopoietic stem cell transplantation: a phase I dose-escalation trial

**DOI:** 10.1101/2021.04.26.21256127

**Authors:** Tessa. M. Andermann, Farnaz Fouladi, Fiona B. Tamburini, Bita Sahaf, Ekaterina Tkachenko, Courtney Greene, Matthew T. Buckley, Erin F. Brooks, Haley Hedlin, Sally Arai, Crystal L. Mackall, David Miklos, Anthony A. Fodor, Andrew R. Rezvani, Ami S. Bhatt

## Abstract

**Background:** Alterations of the gut microbiota after allogeneic hematopoietic cell transplantation (allo-HCT) are a key factor in the development of transplant-related complications such as graft-versus-host disease (GVHD). Interventions that preserve the gut microbiome hold promise to improve HCT-associated morbidity and mortality. Murine models demonstrate that prebiotics such as fructo-oligosaccharides (FOS) may increase gut levels of short-chain fatty acids (SCFAs) such as butyrate, and consequently induce proliferation of immunomodulatory CD4+ FOXP3+ T-regulatory cells (Tregs), that impact GVHD risk.

**Methods:** We conducted a pilot Phase I trial to assess the investigate the maximum tolerated dose of FOS in patients undergoing reduced-intensity (RIC) allo-HCT (n=15) compared to concurrent controls (n=16). We administered FOS starting at pretransplant conditioning and continuing for a total of 21 days. We characterized the gut microbiome using shotgun metagenomic sequencing, measured stool SCFAs using LC-MS, and determined peripheral T-cell concentrations using CyTOF.

**Results:** We found that FOS was safe and well-tolerated at 10g per day without significant adverse effects in patients undergoing reduced-intensity conditioning allo-HCT. Community-level gut microbiota composition was significantly different on the day of transplant (day 0) between patients receiving FOS compared to concurrent controls, however FOS-associated alterations of the gut microbiota were not sustained after transplant. Although the impact of FOS was fleeting, transplantation itself impacted a substantial number of taxa over time. In our small pilot trial, no significant differences were observed in gut microbial metabolic pathways, stool SCFAs, or in peripheral Tregs although Tregs trended higher in those patients who received FOS. Early alterations in gut microbiota composition in those receiving FOS are especially intriguing although it remains unclear what impact this has on outcomes following transplantation and larger studies are required to investigate the use of prebiotics in HCT recipients.

**Conclusions:** FOS is well-tolerated at 10g per day in patients undergoing RIC allo-HCT.

## Introduction

Hematopoietic cell transplantation (HCT) is a lifesaving treatment for patients with hematologic malignancies although success remains limited by disease relapse and infectious or immunologic complications. Innovative supportive care interventions that reduce complications following HCT are essential to decrease treatment-related morbidity and improve non-relapse mortality. The intestinal microbiome has been a major focus of research to improve outcomes following HCT and provides a uniquely modifiable target for both prevention and treatment of HCT-related complications.

The intestinal microbiome functions to maintain intestinal epithelial barrier integrity and immune homeostasis. Perturbations are associated with aberrant local and systemic immune responses that increase the risk of post-transplant complications. Decreased diversity and the presence or absence of specific intestinal organisms are associated with the development of acute graft-versus-host disease (aGVHD) and bloodstream infections^1,2^. The presence of *Blautia* in the gut microbiome is associated with decreased aGVHD and GVHD-related mortality^1^, whereas *Enterococcus* outgrowth in the gut is associated with aGVHD^2^. Additionally, decreased relative abundance of the genera *Faecalibacterium* and *Christensenella* and the family Barnesiellaceae in the gut is associated with increased risk for bloodstream infections^3^. Therefore, interventions that preserve the microbiome are urgently needed to improve HCT-associated morbidity and mortality.

Diet is a key factor impacting gut microbial composition. Indigestible dietary fibers known as prebiotics serve as an important source of nutrition for the community of commensal microbes inhabiting the gastrointestinal tract. Fructo-oligosaccharides (FOS), one of the most well-studied prebiotics, are tolerated up to 20 or 30 g per day in healthy volunteers^4,5^. FOS and other similar prebiotic fibers are thought to support the growth of commensal organisms such as *Bifidobacterium* and to increase levels of gut microbial fermentation products known as short-chain fatty acids (SCFAs). SCFAs, such as butyrate, are thought to induce signaling through chemokine G-protein–coupled receptors, resulting in the proliferation of host CD4+ FOXP3+ regulatory T-cells^6–8^ and the induction of anti-inflammatory cytokines responsible for inhibiting aGVHD in murine models^9^. Prebiotic-induced butyrate also serves as a primary nutritional source for intestinal epithelial cells in addition to its role as histone deacetylase inhibitor, responsible for maintaining intestinal epithelial integrity thought to be essential in preventing GVHD in HCT recipients^4,5,9^. In a small retrospective cohort study of HCT recipients, receipt of a supplement containing 9g/day of FOS in combination with glutamine and fiber (GFO) was associated with significantly less GVHD and improved survival compared to those who did not receive the supplement^10^. A more recent investigation comparing HCT recipients receiving the GFO supplement to historical controls demonstrated a similar decrease in aGVHD with maintenance of gut microbial diversity with supplementation^11^. To date, no studies have comprehensively and prospectively investigated the impact of prebiotic supplementation on SCFAs, Tregs and the gut microbiome in patients undergoing HCT.

In this phase I trial, we use a prospective Bayesian Optimal Interval Design to define maximum tolerated dose, feasibility, and toxicity of FOS for 15 HCT patients undergoing reduced-intensity transplantation. Additionally, we compare the stool microbiome composition, peripheral immune response, and SCFA concentrations of longitudinally collected stool samples from HCT patients who received FOS to contemporaneous controls. In our study, we found that FOS was well-tolerated with only minimal side effects, and intake was primarily limited by oral mucositis. We observed differences in the gut microbiome on the day of transplant between FOS and control groups that were not sustained at subsequent timepoints. Overall, our study suggests that more complex interactions are necessary for sustained effects on gut flora.

## Materials and methods

### Study participants

The current study was a phase I pilot, single-arm, dose-escalation clinical trial performed at Stanford University Hospital over a period of 2 years from 9/13/2016-12/1/2018 (clinicaltrials.gov ID NCT02805075). A total of 15 subjects were recruited to receive fructo-oligosaccharides throughout their transplantation. Stool samples were obtained from all 15 patients at 7 time points throughout the study period (days -5, 0, +7, +14, +28, +60, +100), and blood samples were obtained at day +28 post-transplant. An additional 16 patients undergoing the same reduced-intensity conditioning protocol without receiving FOS also participated in a longitudinal stool collection at the same 7 time points, however blood at day+28 was only available for 5 of these controls. All time points are referenced in the manuscript relative to the day of transplant (day 0). All participants meeting eligibility criteria were informed about the study and required to provide written informed consent under IRB #37979 (PI: Andrew Rezvani) prior to undergoing study-related procedures; patients not receiving FOS had consented to have their stool and blood samples collected under IRB #8903 (PI: David Miklos). The protocol and informed consent documents were reviewed and approved by the Stanford human subjects review board and the Stanford Cancer Institute. The study was performed in accordance with the Declaration of Helsinki.

### Patient eligibility

Adults over 18 years of age with hematologic malignancies undergoing allogeneic HCT as part of a reduced-intensity conditioning protocol were eligible for enrollment. We targeted reduced-intensity allogeneic HCT recipients, reasoning that they would be more likely to tolerate oral intake compared to most myeloablative transplant recipients. No restrictions were made based on underlying malignancy or prior transplant. Patients were excluded if they had undergone gastric bypass surgery, had a history of inflammatory bowel disease, bowel obstruction, known fructose intolerance, or if they were enrolled in a GVHD prevention trial.

### Study design

The study was constructed to find the maximum-tolerated dose using a Bayesian optimal interval study design (see Supplementary Information). The sample size was based on feasibility of accrual within a one-year period. Successive cohorts of participants (3 cohorts with 5 participants per cohort) were enrolled for a total of 15 participants. All 15 patients were to receive one of three possible doses (5g, 10g, or 15g per day) of FOS starting on the day of hospital admission (day -5 prior to transplant) for a total of 21 days. FOS was provided for each patient in the morning as a soluble powder to be dissolved in a beverage or added to food. The dose provided to the second and third groups was dependent on the tolerability of the FOS in the preceding group. We originally defined tolerability as 80% of total doses taken. As this was an arbitrary definition, when our first patient was unable to tolerate FOS due to mucositis, we lowered the threshold of tolerability to 70%. In all subsequent subjects after this first patient, we defined tolerability as the ability to take 70% of total doses.

### Outcomes

Our primary objectives were to determine the maximum tolerated dose of FOS, to describe any adverse events associated with its administration, and to determine the feasibility of providing an oral supplement to patients undergoing reduced-intensity allogeneic HCT. Toxicities were graded using the Common Terminology for Adverse Events Version 4.0 (CTCAE 4.0) and tracked until 100 days after the last dose of FOS^12^. Patients also completed an adverse events survey 14 days following transplant, modeled on the NCI PRO-CTCAE questionnaire for all related gastrointestinal symptoms^13^ (Appendix).

Secondary objectives included correlative analyses of intestinal microbiome and cellular effector response changes in those given FOS compared to controls. Information about clinical outcomes including grades I-IV aGVHD defined according to previously established criteria^14^, bloodstream infection, *Clostridium difficile* infection, and mortality were tracked up to 1 year following transplant.

### Stool and blood collection

Stool samples were collected at 7 time points throughout the study (days -5, 0, +7, +14, +28, +60, +100 relative to transplant) and placed immediately at 4°C upon collection. Within 24 hours, stools were aliquoted into 2mL cryovial tubes and stored at -80°C. Blood samples were collected in heparin collection tubes at day+28 from patients receiving FOS (n=15) and controls (n=5). Peripheral blood mononuclear cells (PBMCs) were isolated using Ficoll (GE Healthcare) density gradient centrifugation according to the manufacturer’s instructions. For long-term storage, PBMCs were resuspended in fetal bovine serum (FBS, Omega Scientific) supplemented with 10% DMSO (Sigma) and stored in liquid nitrogen.

### Metagenomic sequencing of stool samples

DNA was extracted from stool per manufacturer instructions (QIAamp DNA Stool Mini Kit, Qiagen). An additional bead-beating step was added prior to extraction using the Mini-Beadbeater-16 (Biospec Products) and 1-mm diameter zirconia/silica beads (BioSpec Products) and consisting of 7 rounds of alternating 30-s bead-beating bursts followed by 30 s of cooling on ice. The Nextera XT DNA Library Prep Kit (Illumina) was used on DNA extractions to create sequencing libraries. When library preparations were unsuccessful using this method, we instead used the Nextera DNA Flex Library Prep Kit (Illumina) with success in all but 5 samples. Library concentrations were measured using Qubit Fluorometric Quantitation (Life Technologies). Library quality and size distributions were analyzed with the Bioanalyzer 2100 (Agilent). Prepared libraries were multiplexed and subjected to 100 or 150-base pair paired-end sequencing on the HiSeq 4000 platform (Illumina).

### Short-chain fatty acid (SCFA) analysis from stool using LC-MS

Fecal samples were sent to the Metabolomics Core facility at the University of Pennsylvania for SCFA measurement via LC-MS. Samples were homogenized in Volatile Free Acid Mix (5 μL/mg stool, Sigma Aldrich, St. Louis, MO) and centrifuged (13,000g for 5 minutes). The supernatant was filtered using 1.2, 0.65, and 0.22 µm filter plates (Millipore, Billerica, Massachusetts). The filtrate was loaded into Total Recovery Vials (Waters, Milford, MA) for analysis. Short chain fatty acids (SCFAs) were quantified using a Water Acquity UPLC System with a photodiode array detector and an autosampler (192 sample capacity). Samples were analyzed on a HSS T3 1.8 μm 2.1×150 mm column (0.25 mL/min flow, 5 μL injection volume at 40 ^o^C, 25 min per sample run time). Eluent A was 100 mM sodium phosphate monobasic, pH 2.5; eluent B was methanol; the weak needle wash was 0.1% formic acid in water; the strong needle wash was 0.1% formic acid in acetonitrile, and the seal wash was 10% acetonitrile in water. The gradient was 100% eluent A for 5 min, gradient to 70% eluent B from 5-22 min, and then 100% eluent A for 3 min. The photodiode array was set to read absorbance at 215 nm with 4.8 nm resolution. Samples were quantified against standard curves of at least five points run in triplicate. Standard curves were run at the beginning and end of each metabolomics run. Quality control checks (blanks and standards) were run every eight samples. Results were rejected if the standards deviated by greater than ±5%. Concentrations in the samples were calculated as the measured concentration minus the concentration of the solvent; the range of detection was at least 1 – 100 μmol/g stool.

### Immune profiling by mass cytometry

Cryopreserved PBMCs were thawed and washed as described previously^15,16^. Cell-surface antibody master-mix (CSM) (2x) was prepared by adding appropriate dilutions of all cell-surface antibodies into 50 μL CSM per sample. Antibodies targeted were included in the following marker panel: CD3, CD4, CD8, CD11c, CD14, CD16, CD25, CD27, CD33, CD38, CD45, CD45RO, CD45RA, CD56, CD117, CD123, CD127, CD197 (CCR7), TIM3, PD-L1, gdTCR, FcER1a (DCs), TCRVa24-Ja18 (iNKT cells), and HLA-DR. The antibody master-mix was then filtered through a pre-wetted 0.1 μm spin-column (Millipore) to remove antibody aggregates and 50 μL were added to the sample resuspended in 50 μL of CSM. After incubation for 30 min at RT, cells were washed once with CSM. For intracellular staining, cells were fixed using the FoxP3 transcription factor staining buffer set (Thermo Fisher Scientific) to fix and permeabilize for 1h at RT. After fixation, samples were washed once with CSM and once with 1x permeabilization buffer (Thermo Fisher Scientific) and was incubated for 1h with intracellular markers (CD152, FOXP3, Tbet, Ki67). Finally, samples were resuspended in DNA intercalation solution (1.6% PFA (Fisher Scientific) in PBS and 0.5 μM iridium-intercalator (Fluidigm)) for 20 min at RT or overnight at 4°C. Before acquisition, samples were washed once in CSM and twice in ddH_2_O and filtered through a cell strainer (Falcon). Cells were then resuspended at 1 × 10^6^ cells/mL in ddH_2_O supplemented with 1x EQ four element calibration beads (Fluidigm) and acquired on a CyTOF2 mass cytometer (Fluidigm). After acquisition, data from acquired samples was bead-normalized using MATLAB-based software^17^. Barcoded cells were assigned back to their initial samples using MATLAB-based de-barcoding software^18^. Normalized data were uploaded onto the Cytobank analysis platform to perform initial gating and population identification using previously described gating schemes^19^.

### Statistical analyses

All 15 patients receiving FOS and 16 control patients were included in the analyses. The incidence of aGVHD was compared between groups in a cumulative incidence plot using the log-rank test; overall survival was determined for all 31 patients by the Kaplan-Meier method and compared between groups using the log-rank test. Fisher’s exact and Mann Whitney U tests were used for group comparisons of categorical and continuous variables respectively. A p-value less than 0.05 was considered statistically significant based on a two-sided analysis. All p-values were adjusted for multiple hypothesis testing using the Benjamini-Hochberg procedure. Adjusted p-values less than 0.05 were considered significant.

### Computational methods

Stool metagenomic sequencing data were demultiplexed by unique barcodes (bcl2fastq v2.20.0.422, Illumina). Reads were trimmed using TrimGalore v0.5.0^20^, a wrapper for CutAdapt v1.18^21^, with a minimum quality score of 30 for trimming (–q 30) and minimum read length of 60 (–length 60). Trimmed reads were deduplicated to remove PCR and optical duplicates using seqtk rmdup v1.3-r106 with default parameters. Reads aligning to the human genome (hg19) were removed using bwa v0.7.17-r1188^22^. To assess the microbial composition of our short-read sequencing samples, we used the Kraken v2.0.7-beta taxonomic sequence classifier with default parameters^23^ and a comprehensive custom reference database containing all bacterial and archaeal genomes in Genbank assembled to “complete genome,” “chromosome,” or “scaffold” quality as of February 2019. Bracken v2.0.0^24^ was then used to re-estimate relative abundance using Bayesian statistics. Metabolic pathways were characterized using the HUMAnN2 pipeline^25^. The Shannon diversity index was calculated for each sample at the species level using the R package vegan (version 2.5-4). Multidimensional scaling (MDS) with Bray-Curtis distances was performed at each time point to visualize the differences in the microbial composition in FOS and control groups using the “capscale” function from the Vegan package in R. In addition, the ADONIS test was used to compare the microbial composition between FOS and control groups at the species and genus level at baseline and each time point following transplant. The Student’s t-test was used to compare changes in the relative abundances of species at each time point (0,7,14,28,60, and100 days) relative to baseline (day -5) between FOS and control groups. For this analysis, only patients having fecal samples at both the baseline (day -5) and timepoint after transplant were included. (day 0: control=6 FOS=14; day 7: control=7 FOS=13; day 14 control=7 FOS=13; day 28 control = 7 FOS 13; day 60 control=8 FOS=12; day 100 control=7 FOS=12). To model overall changes in the gut microbiome following transplant, mixed linear models with a second-degree polynomial for time as a fixed effect and patient ID as a random effect *(taxa ∼ -day*^*2*^, *random = ∼1* | *patient ID)* were constructed using the “lme” function from the nlme package in R. The polynomial term in these models allowed us to model an initial decrease and then increase in the log_10_ relative abundances of taxa following transplant. The Akaike information criterion (AIC) was used to compare the fitting of the model with alternatives that did not include a polynomial *(taxa ∼ -day, random = ∼1* | *patient ID)*, include a third- or fourth-degree polynomial (taxa ∼ -day^3 or 4^, random = ∼1 | patient ID), and include the FOS term in the model *(taxa ∼ -day*^*2*^ *+ FOS, random = ∼1* | *patient ID)*. Only taxa that were present in more than 25% of samples were included in the analysis. We chose this second order model for simplicity and power. While it does make strong parametric assumptions, the overall magnitude of changes seen with time have strong enough effect sizes so that significant results are robust to the assumptions. We have previously used these models to fit changes in microbial abundance over time^26^. All p-values were adjusted for multiple hypothesis testing using the Benjamini-Hochberg procedure. Adjusted p-values less than 0.05 were considered significant. Similar analyses were performed to compare the metabolic pathways characterized by the HUMAnN2 pipeline between FOS and control groups following transplant.

Plots were generated in the R programming language (v3.5.2) using the following packages: ggplot2 v3.1.1, genefilter v1.64.0, RColorBrewer v1.1-2, ggpubr v0.2, cowplot v0.9.4, scales v1.0.0, gtools v3.8.1, reshape2_1.4.3, dplyr_0.8.0.1, plyr_1.8.4, ggplot2_3.1.1.

## Results

### Patient characteristics

A total of 16 individuals undergoing HCT at Stanford were enrolled in the study and received FOS between 9/2016 and 10/2017 (Table 1). One study subject dropped out prior to starting the intervention; this individual was replaced with an additional enrollee and thus a total of 15 subjects completed the study. All study subjects underwent reduced-intensity allogeneic HCT with Fludarabine plus Melphalan conditioning and FK506/Methotrexate GVHD prophylaxis. All patients undergoing allo-HCT at Stanford received oral Ciprofloxacin for gut decontamination starting on the day of conditioning and continuing until engraftment. As part of the standard protocol, acyclovir and fluconazole are started on day +1 unless the patient is already on a mold-active antifungal agent. Trimethoprim-sulfamethoxazole is started for *Pneumocystis jiroveci* prophylaxis at day +42. All other medications including antibiotics were given at the discretion of the treating physician. Stool and blood samples were collected from 15 patients receiving FOS and from 16 control patients undergoing the same reduced-intensity protocol between 9/2016-9/2018 who did not receive FOS (Table 1).

**Table 1.** Patient characteristics did not significantly differ between those receiving FOS compared to controls (Wilcoxon rank sum and Fisher’s exact test, p < 0.05).

|  | FOS (n=15) | Controls (n=16) | p-value |
| --- | --- | --- | --- |
| Mean age (years) | 61 | 65 | 0.095 |
| Sex (%) |  |  |  |
| Female | 7 (47%) | 9 (56%) | 0.72 |
| Race (n, %) |  |  |  |
| White | 8 (53%) | 11 (69%) | 0.47 |
| Asian | 3 (20%) | 2 (13%) | 0.65 |
| Black | 2 (13%) | 1 (6%) | 0.6 |
| Pacific Islander | 0 (0) | 0 (0) | 1 |
| Unknown | 2 (13%) | 2 (13%) | 1 |
| Ethnicity (%) |  |  |  |
| Hispanic | 1 (7%) | 0 (0) | 0.48 |
| Underlying malignancy |  |  |  |
| Acute myelogenous leukemia | 5 (33%) | 6 (38%) | 1 |
| Non-Hodgkin lymphoma | 3 (20%) | 1 (6%) | 0.33 |
| Myelodysplastic syndrome | 5 (33%) | 7 (44%) | 0.72 |
| Chronic myelogenous leukemia | 1 (7%) | 1 (6%) | 1 |
| Blastic plasmacytoid dendritic cell neoplasm | 1 (7%) | 1 (6%) | 1 |
| Donor type |  |  |  |
| Matched related | 7 (47%) | 4 (25%) | 0.27 |
| Matched unrelated | 7 (47%) | 9 (56%) | 0.72 |
| Mismatched unrelated | 1 (7%) | 3 (19%) | 0.6 |
| Donor product |  |  |  |
| Peripheral blood | 15 (100%) | 14 (88%) | 0.48 |
| Bone marrow | 0 (0) | 2 (13%) |  |
| Antibiotics |  |  |  |
| Non-carbapenem beta-lactams | 12 (80%) | 9 (56%) | 0.25 |
| Carbapenems | 3 (20%) | 5 (31%) | 0.69 |
| IV Vancomycin | 9 (60%) | 6 (38%) | 0.29 |

Demographic and clinical characteristics of the intervention and control patients shown in Table 1 demonstrate that there were no significant differences between patients in the two groups.

### FOS was well-tolerated

The primary goal of the study was to determine the maximum-tolerated dose of FOS in patients undergoing allogeneic HCT. The trial was constructed using the Bayesian Optimal Interval (BOIN) dose-escalation trial design with 3 groups of 5 patients receiving increasing doses of FOS depending on the tolerability of the prior group (see Methods and Supplementary Information for details). The first group of 5 patients received a mean of 80% of total doses over 21 days, and 4 of 5 patients in this group met criteria for tolerability. The next group of 5 patients received an increased dose of 10g of FOS per day. While the mean intake of this second group remained at 80% of total doses, only 3 of 5 patients met criteria for FOS tolerability and we were unable to advance the third and final group of 5 patients to the highest dose (15g). The third group on average took 71% of total doses. We therefore found our maximum-tolerated dose in this study to be 10g of FOS per day. Intake of FOS was limited in this group by significant chemotherapy-induced nausea and mucositis.

### Adverse events were primarily due to regimen-related toxicity

In total, 160 adverse events in patients receiving FOS were identified, with 14 reported to be possibly related to FOS and the remaining thought unlikely or unrelated to the prebiotic (Supplementary Table 1). A total of 6 patients experienced severe adverse events (SAEs) including neutropenic fever (2), disease relapse (1), severe pain (1), delirium (1), and severe mucositis (1). The top 10 most reported episodes of AEs were nausea (n=17), diarrhea (n=16), and rash (n=14), mucositis (n=12), lower extremity or pedal edema (n=11), vomiting (n=10), non-abdominal pain (n=8), abdominal pain (n=7), fever (n=7), and abdominal distension (n=5). Adverse events that were reported as “possibly related” include bloating (n=5), abdominal pain (n=4), flatulence (n=4), and constipation (n=1). All of these were reported as mild or moderate and in only 2 cases did these symptoms lead to discontinuation of FOS. Otherwise, no action was taken, or the FOS was temporarily discontinued (n=2) and restarted. No adverse events were reported as clearly attributable to FOS. Patient reported outcomes for gastrointestinal symptoms were also assessed at day+14 in those receiving FOS and were not higher in patients receiving 10g per day compared to those receiving 5g per day (Supplementary Table 2).

### Clinical outcomes did not differ between groups

FOS and other carbohydrate-based prebiotics are fermented by commensal gut bacteria to produce short-chain fatty acids (SCFAs). Not only are SCFAs are an important source of energy for intestinal epithelial cells and the maintenance of an intact intestinal mucosal barrier, they have been demonstrated to impact immune intestinal homeostasis and increase the proliferation of T-regulatory cells, important cellular effectors for the attenuation of inflammation in the gut^27^. We hypothesized that prebiotics may be helpful in decreasing aGVHD risk and therefore reducing overall mortality. We found that neither aGVHD (grades I-IV) or overall survival at 1 year were impacted by the administration of FOS compared to controls (Table 2, Figures 1a and 1b).

**Table 2.**
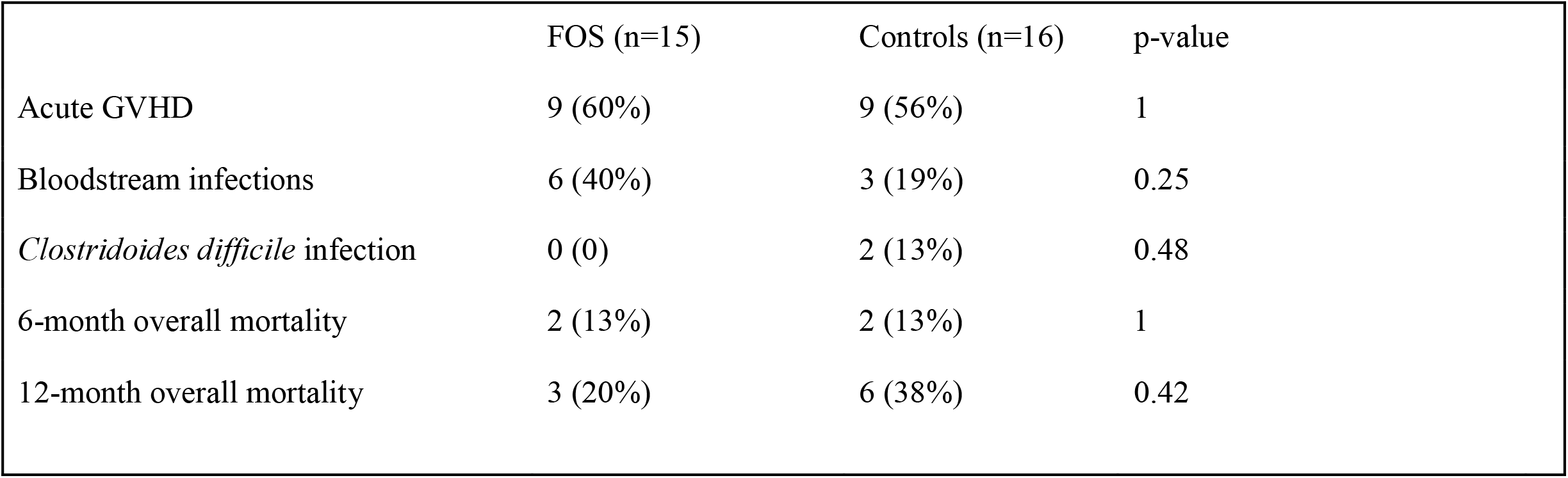
Transplant outcomes do not significantly differ between patients who received FOS compared to controls. FOS was not associated with significant differences in acute graft-versus-host disease (GVHD), bloodstream infection, *Clostridioides difficile* infection, or 6-month and 12-month mortality compared to control patients (Fisher’s exact test, p < 0.05).

|  | FOS (n=15) | Controls (n=16) | p-value |
| --- | --- | --- | --- |
| Acute GVHD | 9 (60%) | 9 (56%) | 1 |
| Bloodstream infections | 6 (40%) | 3 (19%) | 0.25 |
| *Clostridioides difficile* infection | 0 (0) | 2 (13%) | 0.48 |
| 6-month overall mortality | 2 (13%) | 2 (13%) | 1 |
| 12-month overall mortality | 3 (20%) | 6 (38%) | 0.42 |

**Figure 1a.**
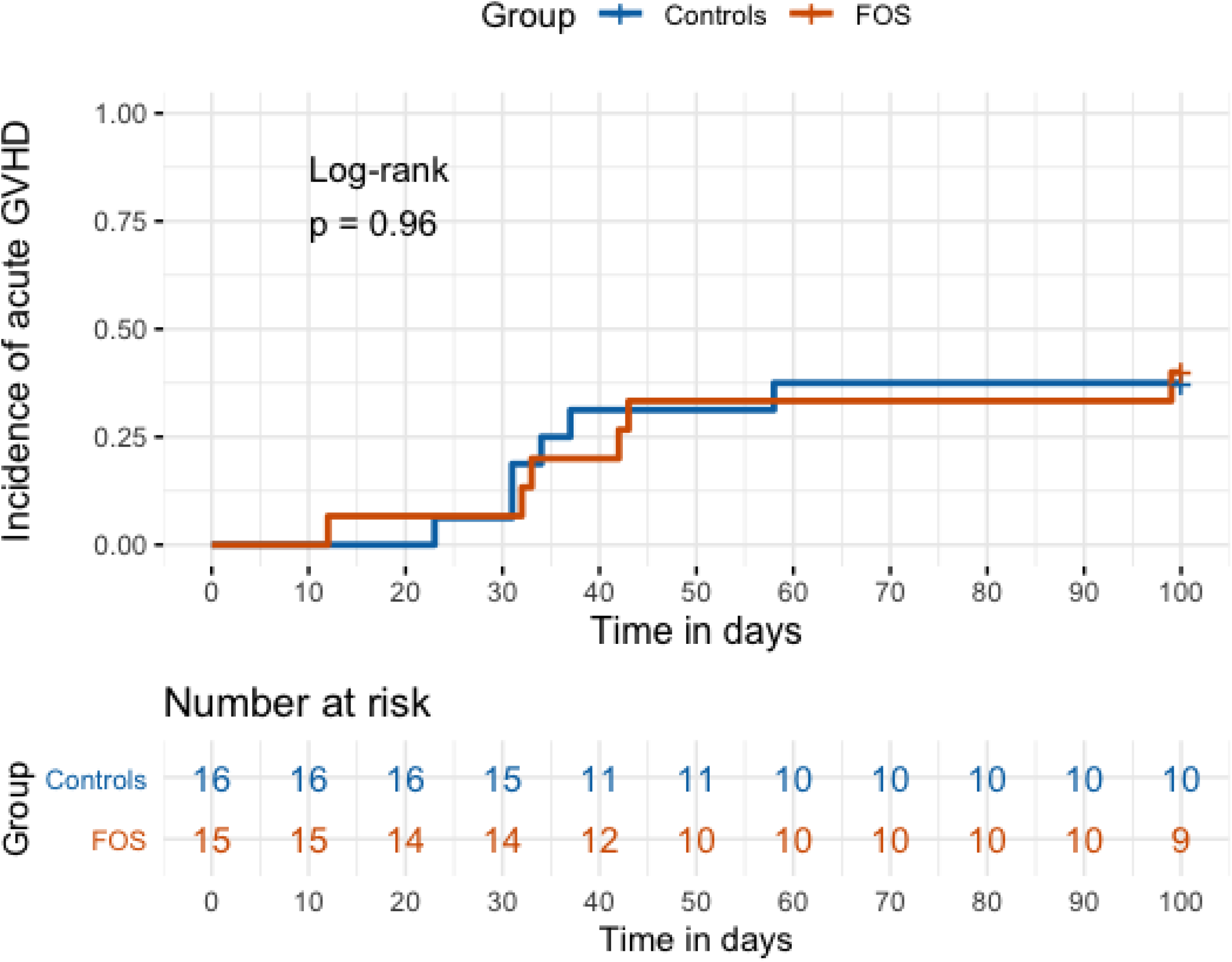
Cumulative incidence analysis shows no significant difference in acute GVHD between groups. Cumulative incidence of grades I-IV acute graft-versus-host disease (GVHD) was compared between those who received FOS and contemporaneous controls. Patients who received FOS were no different in their risk of acute GVHD compared to control patients who did not receive FOS (log-rank test, p=0.96).

**Figure 1b.**
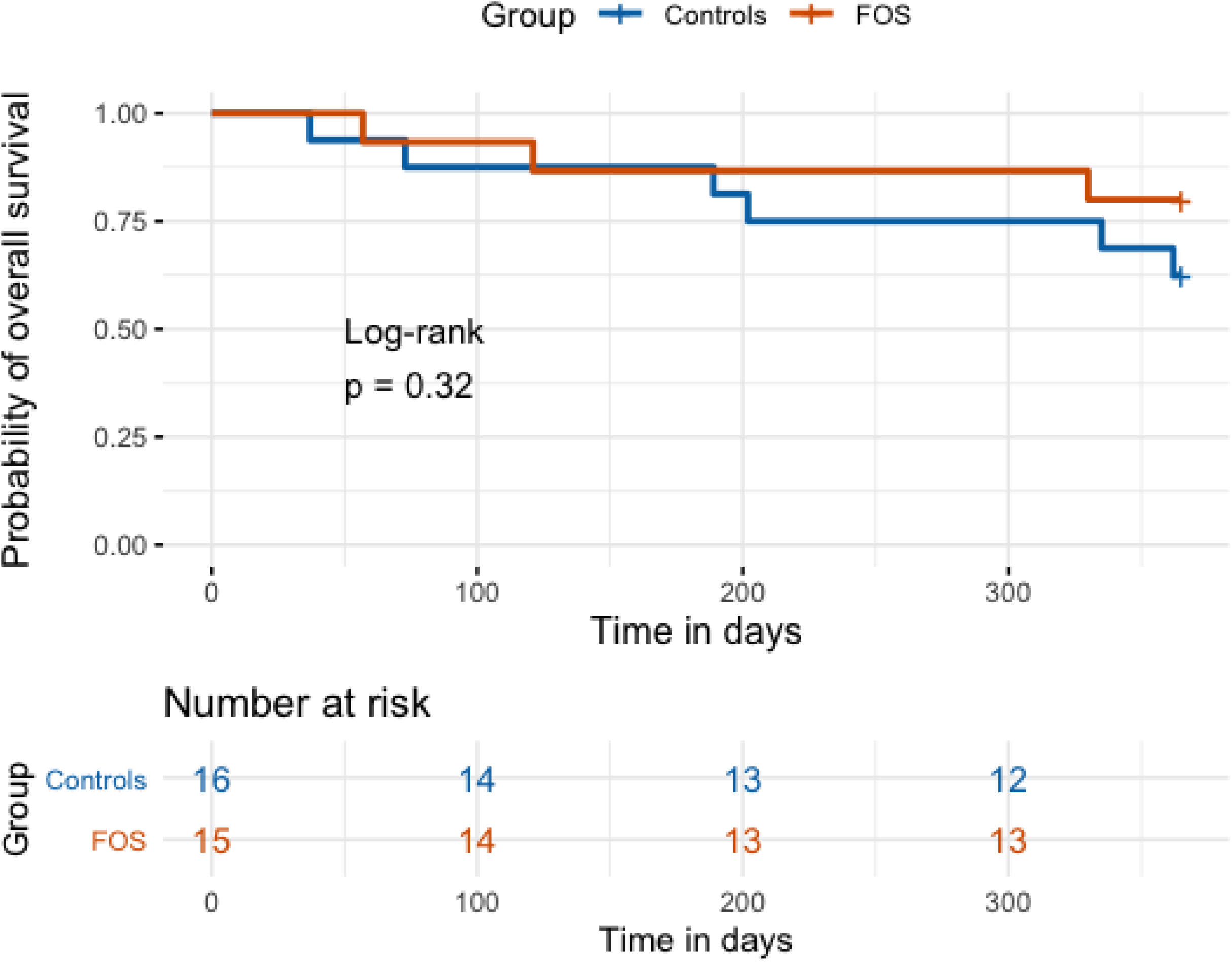
Kaplan-Meier analysis demonstrated no significant difference in survival between patients who received FOS compared to controls at 1 year. Overall survival at 1 year was compared between those who received FOS and contemporaneous controls. Patients who received FOS were no different in their overall survival compared to control patients who did not receive FOS (log-rank test, p=0.32).

Patients were monitored for infectious outcomes for 100 days post-transplant. Both *Clostridium difficile* associated colitis (CDAD) and bloodstream infections (BSI) result from an abnormal, low diversity microbiome lacking in obligate anaerobic bacteria^28–30^. We compared the incidence of CDAD and BSI in patients receiving FOS to controls and found that while CDAD occurred in only 2 controls and no FOS patients, twice as many patients receiving FOS developed a bloodstream infection (40% FOS v. 19% controls) although this difference was not statistically significant (Table 2). All 9 BSIs recorded for both groups occurred with organisms thought to originate from the oral or gastrointestinal microbiome such as Enterobacteriaceae, *Streptococcus mitis*, and *Enterococcus* species.

### Gut microbiomes in patients receiving FOS were similar to control patients

Prior studies have demonstrated higher levels of specific commensal bacteria, including *Bifidobacterium spp*. following administration of FOS in healthy volunteers^31^. We hypothesized that FOS would alter the gut microbiome and that patients receiving FOS would have higher Shannon diversity at day +14. Stool samples at 7 times points in both FOS and control patients underwent DNA extraction, shotgun metagenomic sequencing, and taxonomic classification (see Methods). The number of sequenced raw reads per sample was substantially reduced following quality filtering and exclusion of human reads (median number of 28,083,873 raw reads v. 8,129,375 filtered reads per sample: Supplementary Table 3, Supplementary Figure 1).

We calculated Shannon diversity, a measure of microbial species richness and evenness, for each stool and compared measurements over time. In both groups, diversity declined immediately following transplant and returned to near baseline levels at day+100 (Figure 2a). No significant differences in diversity were observed between patients receiving FOS compared to controls at any of the 7 time points (Figure 2b).

**Figure 2.**
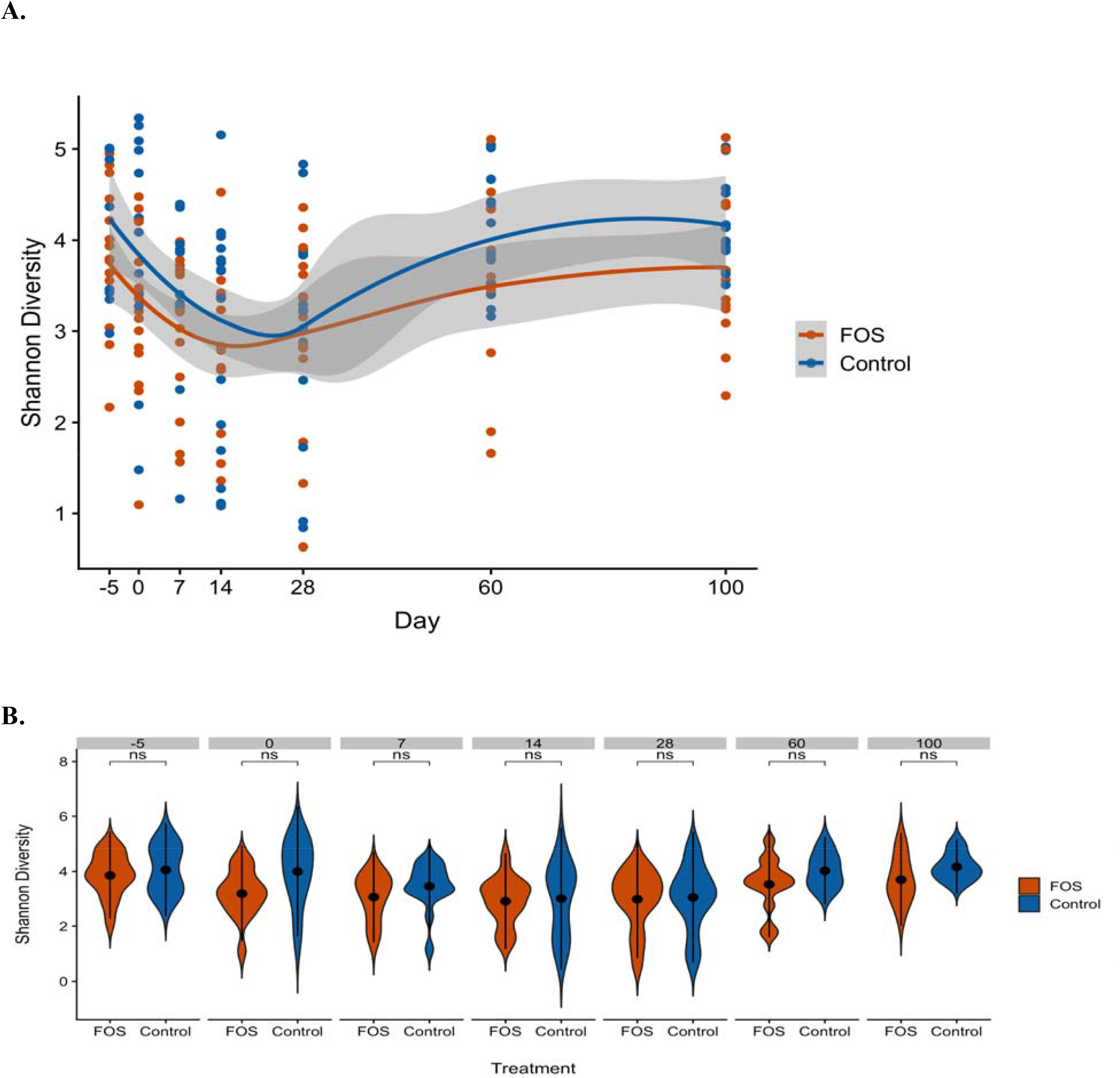
FOS treatment is not associated with changes in alpha diversity relative to controls. Shannon diversity, a measure of species richness and evenness, was calculated for each microbiome sample. (**A**) Shannon diversity declines after transplant and is restored by day 100 post-transplant. Regression lines were fit using LOESS smoothing, with gray bands representing 95% confidence intervals. (**B**) Shannon diversity is not significantly different between FOS-treated patients and controls at any timepoint (Wilcoxon rank sum test, FDR-adjusted q < 0.05).

To test the hypothesis that FOS treatment shifted the microbial community, we analyzed longitudinal whole-genome shotgun metagenomic sequences from 15 subject given FOS treatment and 16 control subjects. Multidimensional scaling (MDS) of stool taxonomic composition revealed a distinct cluster between FOS and control only at time-point zero (ADONIS test, R^2^=0.07, p-value=0.03), 5 days after initiation of FOS, but not at baseline or at later timepoints (Figure 3). At day 0, the day of transplant, FOS and control groups were significantly associated with the second axis of the ordination plot, which explains 12.2% of the variation at this timepoint (Simple linear regression, R^2^=0.31, adjusted p = 0.03). No other timepoints were significantly different between FOS and control for either of the first two MDS axes (Supplementary Figure 2).

**Figure 3.**
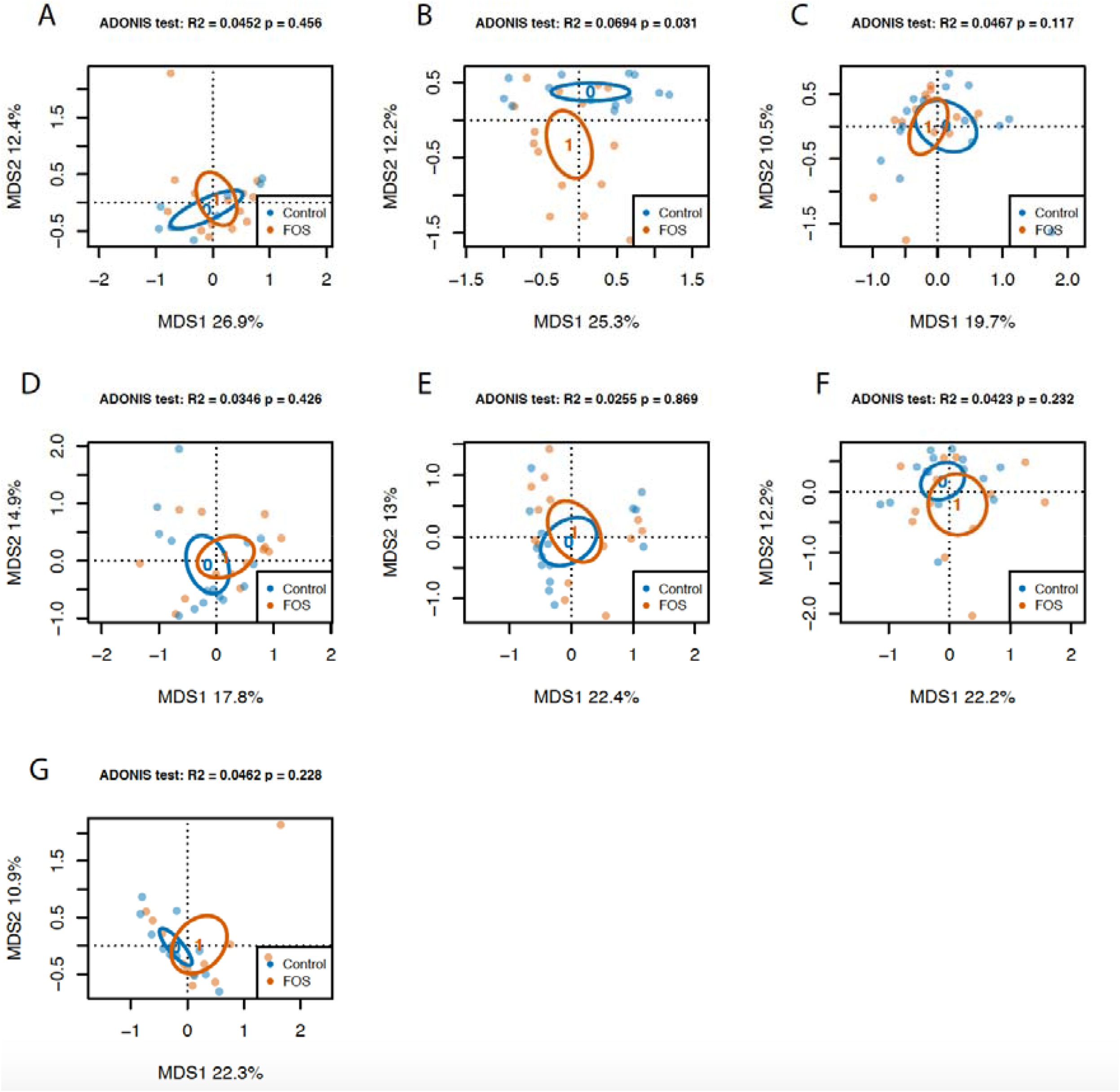
Gut microbiota composition is significantly different between FOS and Control groups only at day 0, on the day of transplantation. Multidimensional scaling (MDS) of gut microbiota taxonomic composition at the species level were performed using Bray-Curtis distances at each time point relative to transplantation (A: day -5, B : day 0, C: day 7, D: day 14, E: day 28, F: day 60, G: day 100). The ADONIS test was used to compare the gut microbiota composition between FOS and control groups at each time point.

Our multidimensional scaling (MDS) data are consistent with FOS treatment producing a transient response in the microbial community. To provide a more detailed view of microbial composition following FOS intervention, we tested the hypothesis that the microbial community responds differently to transplant in FOS group versus the control group. For this, we compared changes in in the relative abundances of individual taxa at each timepoint relative to baseline (−5 day) using a Student’s t-test. We found no significant differences in the response to transplant between FOS and control groups at any timepoints following transplant (Supplementary Figure 3). The fact that we saw a difference in the MDS plot at day 0 between FOS and control groups but not a significant difference in any individual taxa may reflect the small sample size and the potential rejection of true positives after correction for multiple hypothesis testing.

Since we observed both FOS and control groups respond similarly to transplant, we next examined the effect of transplant on the log_10_ relative abundance of taxa regardless of the FOS intervention. Similar to the pattern we observed for Shannon diversity (Figure 2), for many taxa, we observed a decrease in abundance following transplant and then a recovery (Figure 4). To model this, we used mixed linear models with the second-degree polynomial for time as a fixed effect and patient ID as a random effect. AIC analysis indicated that this model was a reasonable fit compared to alternatives that included terms for FOS or did not include a polynomial term or had a third-degree or fourth-degree polynomial (Supplementary Figure 4). According to this model, 3,081 out of 13,308 species underwent significant changes over time following transplant (Supplementary Table 4) at a 5% FDR threshold. Taxa with the most changes significant changes following transplant included *Ruminococcus spp*., *Clostridium spp*., and *Enterobacteriaceae bacterium* (Figure 4).

**Figure 4.**
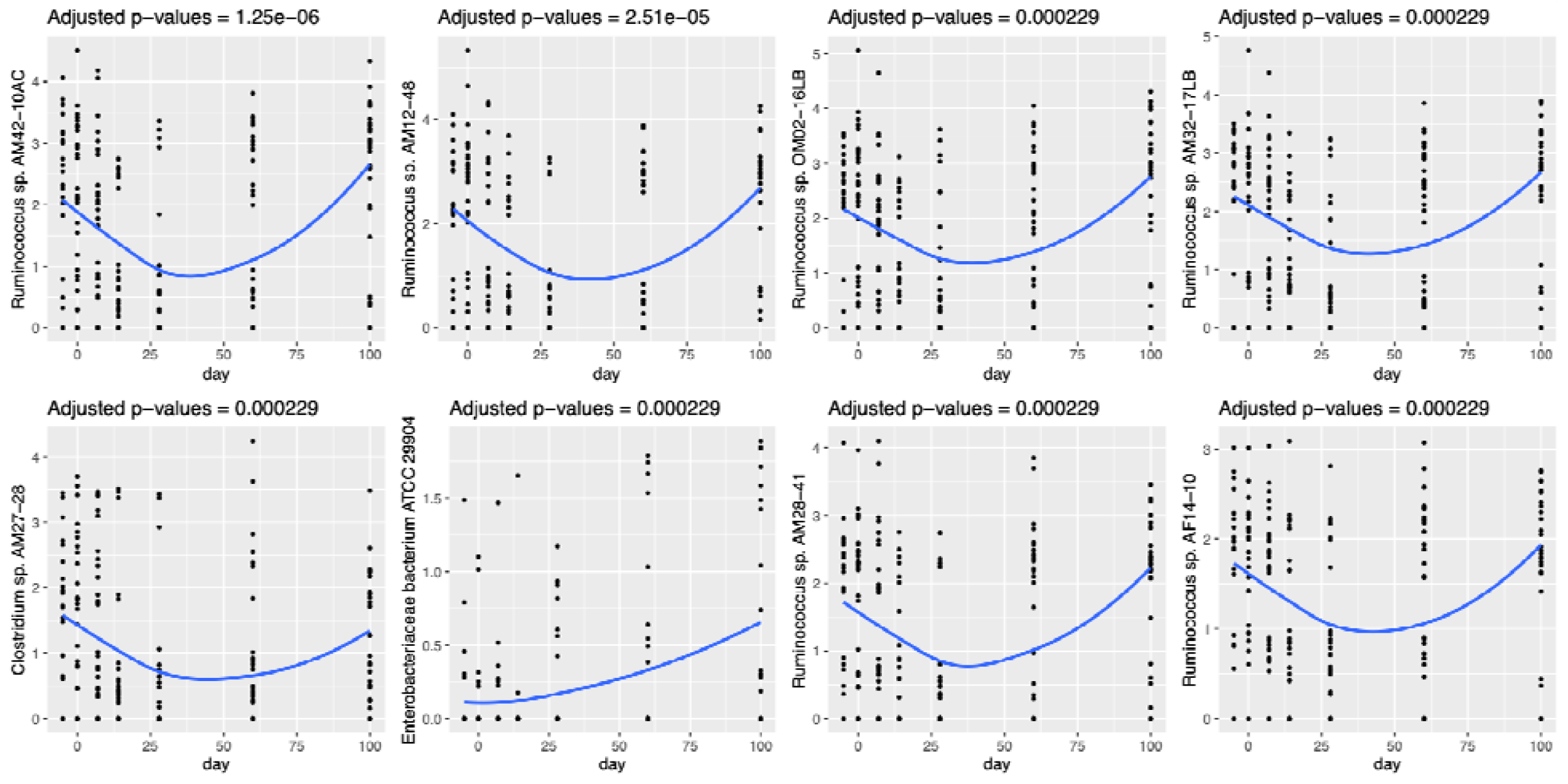
The gut microbiota composition undergoes significant changes following transplant. The plots show the relative abundance of the most significant taxa from mixed linear models with a second-degree polynomial for time as a fixed effect and patient ID as a random effect (taxa ∼ - day^2^, random = ∼ 1 | patient ID). The y-axis represents the log_10_ relative abundance of taxa. The blue line shows the predicted values for the log_10_ relative abundance of taxa at each time point using the mixed linear model.

We next examined the abundances of metabolic pathways produced by the HUMAnN2 pipeline of the gut microbiome at each timepoint following transplant. Unlike the taxonomic composition of the gut microbiota, the relative abundances of metabolic pathways were not different between FOS and control groups at any timepoint (Supplementary Figure 5) and changes in metabolic pathway abundances were not different between FOS and control groups. Mixed linear models with a second-degree polynomial for time as a fixed effect and patient ID as a random effect revealed that the relative abundance of pathways changed following transplant (Supplemental Figure 6). 146 out of 311 pathways were significantly altered over time at a 5% FDR. Examples of pathways with significant changes following transplant include the super pathway of N-acetylneuraminate degradation, the super pathway of histidine, purine, and pyrimidine biosynthesis, glycerol degradation to butanol, the super pathway of glyoxylate cycle and fatty acid degradation, and the super pathway of hexitol degradation (Supplementary Table 5). In many cases, metabolic pathway relative abundance dips briefly after transplant and increases post-transplant to levels higher than at baseline.

### SCFAs did not differ between patients based on FOS intake

We hypothesized that those patients receiving FOS would have increased levels of SCFAs in their stool potentially due to enrichment in organisms that ferment FOS to produce SCFAs such as butyrate. Unsurprisingly, given the lack of sustained enrichment of microbial taxa over time, stools analyzed using LC-MS at day +14 did not demonstrate significant differences in levels of butyrate, propionate, or acetate (Figure 5).

**Figure 5.**
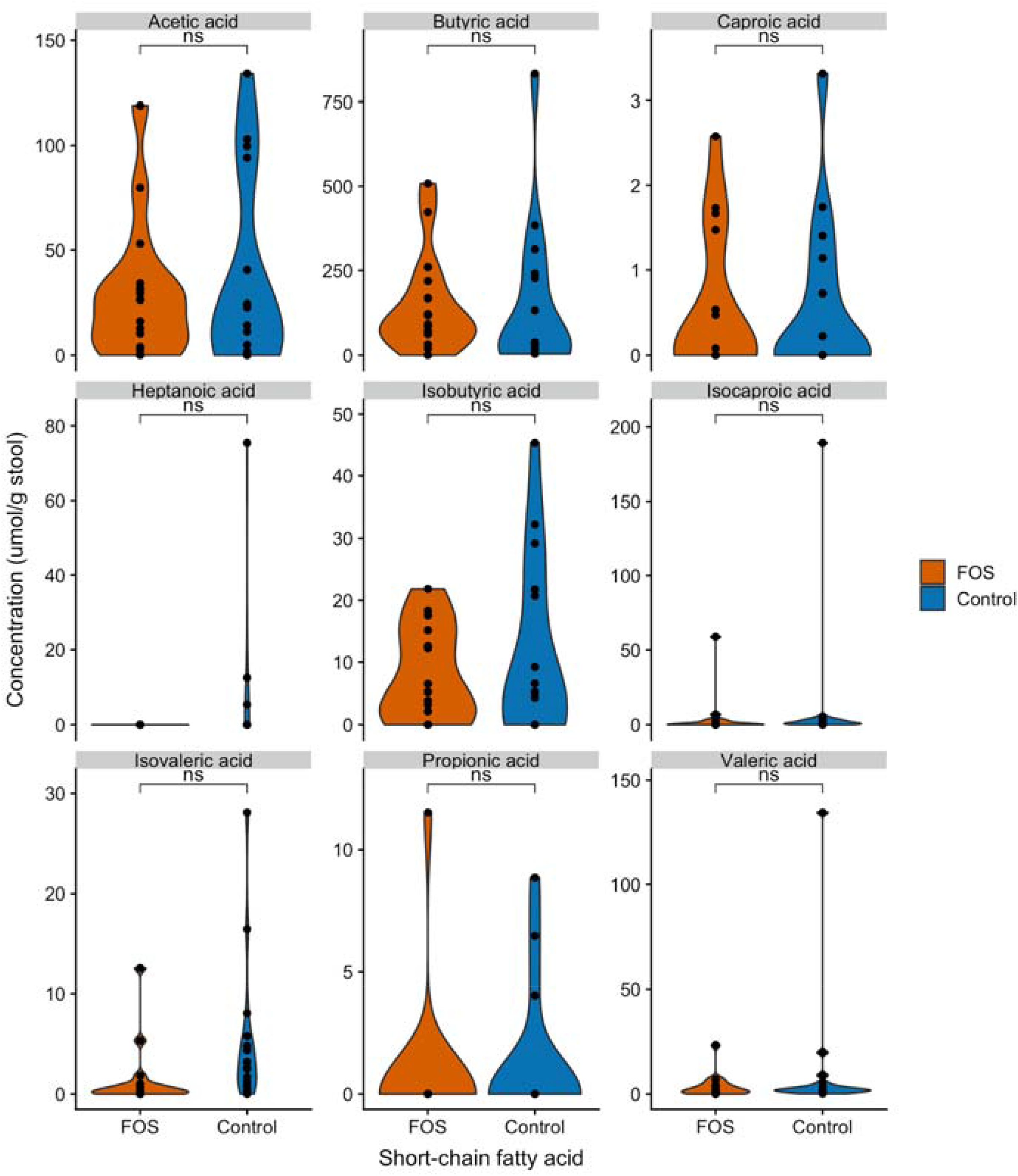
Short-chain fatty acid concentration in stool is not different between FOS-treated and control patients. Short-chain fatty acids were measured using mass spectrometry (LC-MS). No significant differences were detected in short-chain fatty acid concentration between patients treated with FOS and controls (Wilcoxon rank sum test, FDR-adjusted q < 0.05).

### CyTOF of peripheral blood demonstrates similar immune profiles between FOS-treated patients and controls

Microbially-produced short-chain fatty acids impact host immune response in part through induction of intestinal regulatory T cells (Tregs). SCFAs such as butyrate induce the colonic differentiation of CD4+FOXP3+Tregs, crucial to host intestinal immune homeostasis^6,7^. To determine whether FOS intake impacted concentrations of peripheral Tregs after transplantation, blood samples were obtained from 15 patients who received FOS and 5 control patients to determine whether FOS altered peripheral immune reconstitution at day+28 following hematopoietic stem cell transplantation compared to controls. Whole-blood derived PBMCs were subjected to a previously validated, comprehensive CyTOF workflow developed for identifying immune signatures for GVHD in peripheral blood following HCT^15,16^. Complete results comparing median absolute cell concentrations between FOS and control groups are shown in Supplementary Figure 7 with unadjusted p-values. Although median concentrations of CD4+FOXP3+Tregs differed to a great extent between FOS-treated patients and controls (2.1 v 0.6 cells/ μL), this difference was not statistically significant even before adjusting for multiple hypothesis testing (unadjusted p=0.07). Other immune responses following transplantation were also similarly not statistically significant after adjustment for multiple comparisons (Supplementary Figure 7).

## Discussion

The microbiome has emerged as a promising potential contributor to allo-HCT patient outcomes. However, to date, few prospective interventional trials have investigated the impact of microbiome-modifying therapies (e.g., diet, antibiotics, prebiotics, probiotics) for their potential impact on outcomes such as aGVHD incidence and overall survival. In this study, we performed a prospective trial of prebiotics in HCT recipients with concurrent controls. We found that FOS was well-tolerated at a maximum-tolerated dose of 10g per day, limited only by chemotherapy-induced nausea and mucositis. Patients reported mild abdominal distension and flatulence, and in only two patients were these effects dose-limiting. No significant differences in adverse outcomes including GVHD, infection, or mortality were observed in association with FOS administration, although our study was underpowered to detect small differences.

In our pilot, we found that FOS was associated with community-level taxonomic differences on the day of transplant (day 0). These differences between groups did not endure beyond transplantation, perhaps as a result of the variability in antibiotic use that occurs with febrile neutropenia treatment. Following day 0, a profound change in the microbial community was observed, consistent with prior literature ^32^; as many as 20% of the taxa were altered while often recovering to pre-transplant levels, albeit slowly over 100 days (Figures 2 and 4). In addition to a modest short-term microbial signal associated with FOS intake, there was a non-significant trend of higher peripheral FOXP3+ T-regulatory cells in those who received FOS compared to controls, in line with prior murine models^33^. Although we would have expected differences in stool SCFAs between groups, none were observed at day 28. These data demonstrate that any signal observed as a result of FOS was temporary and likely overwhelmed by other factors, including antibiotics, and dietary intake following transplantation.

The limitations of our study are multiple, including that we performed our investigation at a single center. As our study was not powered to detect differences between groups resulting from FOS intake, our ability to observe differences in taxonomic composition and clinical outcomes was greatly limited due to sample size. In addition, FOS was not taken consistently throughout the post-transplant period due to severe mucositis. Intermittent or truncated prebiotic intake may have impacted our ability to detect changes in microbiome composition or clinical outcomes compared to prior studies demonstrating significant prebiotic efficacy, in which adherence was more consistent and FOS was administered for a longer duration both pre- and post-transplantation (21 v. 35 days)^10,11^. Antibiotics administered before transplant may have also eliminated many of the bacteria capable of using FOS for nutrition. Indeed, Yoshifuji and colleagues demonstrated that patients with a higher baseline microbiome diversity appeared more likely to benefit from prebiotic supplementation, potentially due to the greater presence of prebiotic fermenting bacteria^11^. In a prior study in which FOS intake was associated with decreased rates of aGVHD^10^, patients not only received FOS, they also received a *Lactobacillus* probiotic as part of standard of care. The use of a “synbiotic”, combining a prebiotic with a probiotic, may be more efficacious in improving clinical outcomes and positively impacting the microbiome compared to using a prebiotic alone, as demonstrated in other gastrointestinal disorders such as inflammatory bowel disease^34^. In both prior published studies investigating FOS use in HCT recipients^10,11^, supplementation of dietary fibers with resistant starches in combination with oligosaccharides may have been more effective at maintaining gut microbial diversity and improving clinical outcomes. Longer duration of prebiotic use both before and after HCT along with multi-fiber supplementation may therefore demonstrate greater prebiotic efficacy and more significant, durable maintenance of the baseline microbiota throughout transplantation.

Although our findings demonstrate a limited community-level microbial signal on the day of transplant associated with FOS intake, the pre- and peri-transplant microbiome has demonstrated significant associations with favorable outcomes following HCT^30,35^. To maintain a greater proportion of the baseline microbiota, further investigation of pre-emptive microbiota-targeting therapies given for a longer duration prior to transplantation or conditioning in larger randomized studies is worthwhile. Additionally, therapies targeting a greater number of microbes may require more than one prebiotic at a time with or without a probiotic/synbiotic, capable of maintaining microbiome diversity and essential commensal bacteria throughout transplant. It may be beneficial to determine whether organisms capable of fermenting specific prebiotics are present in the pre-transplant gut microbiome before patients undergo allo-HCT. For patients lacking the necessary commensals, personalized synbiotic cocktails compromised of SCFA-producing bacteria maintained via prebiotic administration may be required to supplement baseline gut flora prior to transplant. Personalization of microbiome-altering therapies and supplementation of the gut microbiome prior to transplant -in combination with judicious antibiotic use to maintain baseline microbiota may offer a unique and promising method of mitigating complications from stem cell transplantation and improving survival outcomes

## Data Availability

Data not included in the manuscript will be available by request to the corresponding authors. All metagenomic data will be uploaded to the SRA.

## Acknowledgements

We would like to thank the members of the Bhatt Lab for providing feedback on the study design, the bioinformatics pipeline, and manuscript revisions. We especially thank the patients and nurses on the Blood and Marrow Transplantation service for their enthusiastic participation in this project. This work was supported in part by the Amy Strelzer Manasevit Award from the National Marrow Donor Program, the National Center for Advancing Translational Science, Clinical and Translational Science Awards KL2TR001083 and UL1TR001085 and the Society of Blood and Marrow Transplantation New Investigator Award (T.M.A.). F.B.T was supported in part by the National Science Foundation Graduate Research Fellowship. A.S.B. was supported by NCI KO8 CA184420, the Amy Strelzer Manasevit Award, a Damon Runyon Clinical Investigator Award, and the V Foundation scholar award.

