## Supplementary figures for "A fructo-oligosaccharide prebiotic is well-tolerated in adults undergoing allogeneic hematopoietic stem cell transplantation: a phase I dose-escalation trial"

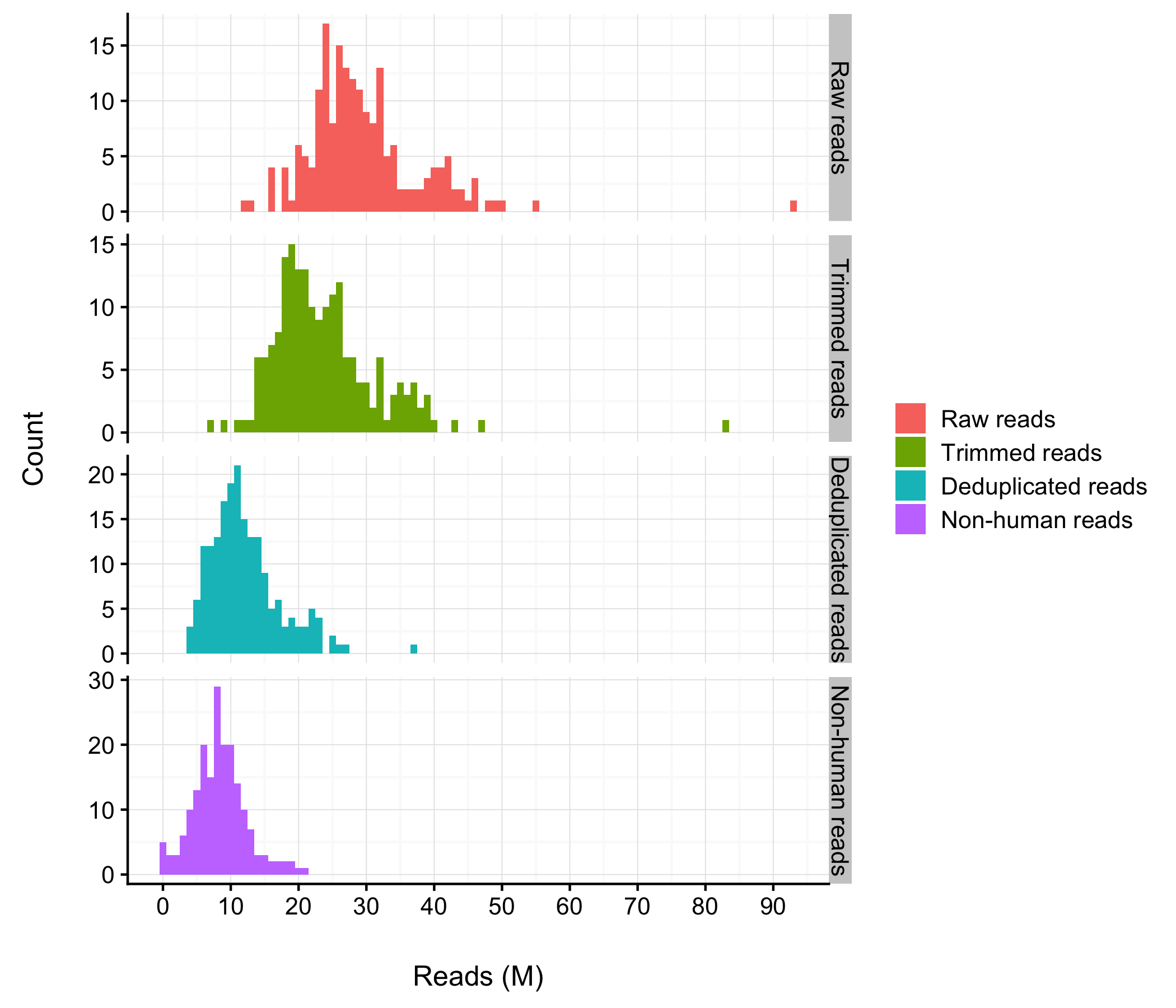

**Supplementary Figure 1. Quality filtering of sequenced reads.** The number of sequenced reads per sample was substantially reduced following quality filtering, including trimming of adapter sequences, deduplication of sequences, and exclusion of human reads.

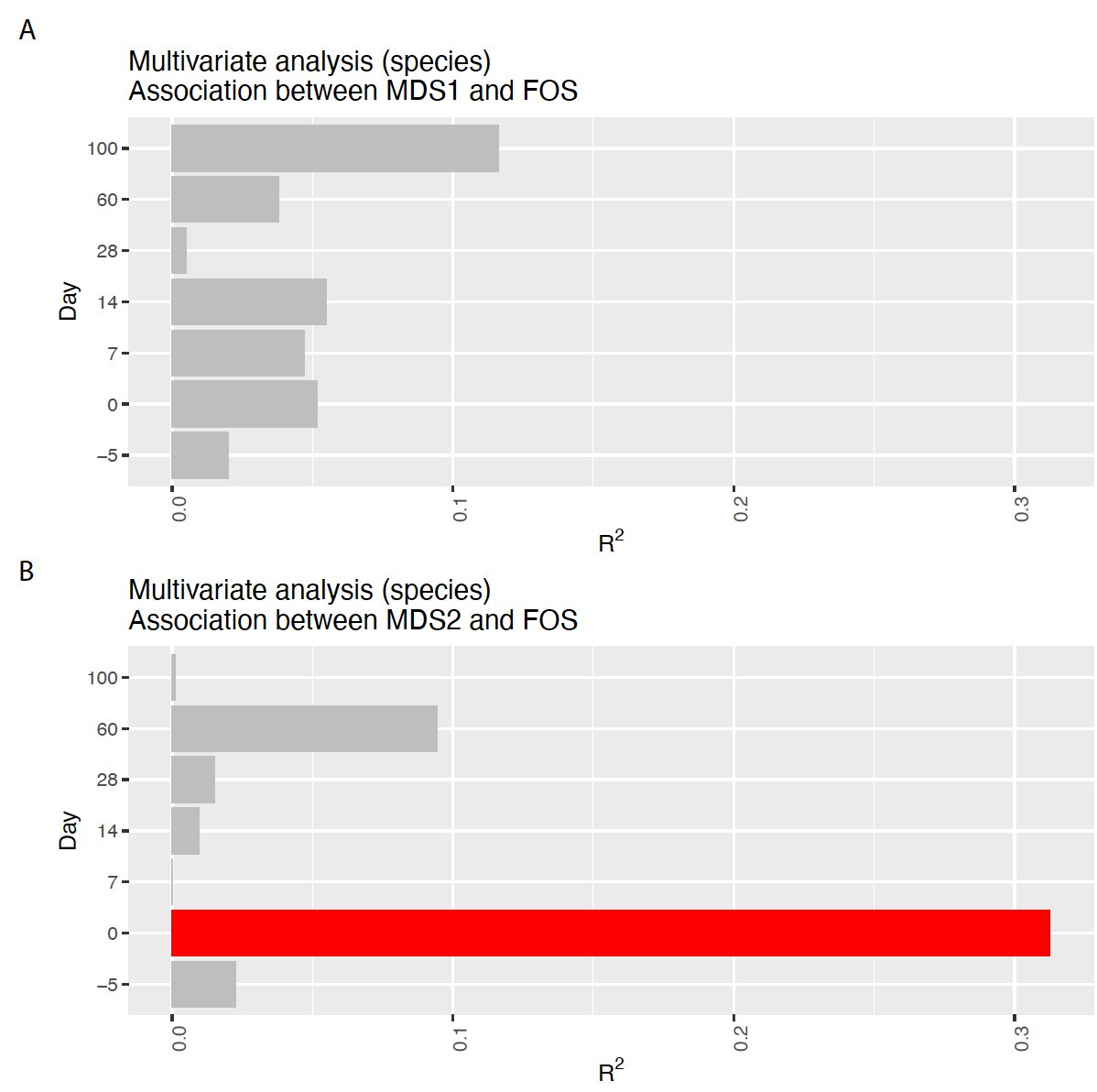

**Supplementary Figure 2. Gut microbiota composition at day 0 is associated with FOS intake compared to controls.** The effect size (R^2^) from simple linear regression models with the first (A) and second (B) PCoA axes at each timepoint as dependent variables and FOS versus control as an independent variable. Red bars indicate statistical significance at FDR 5%.

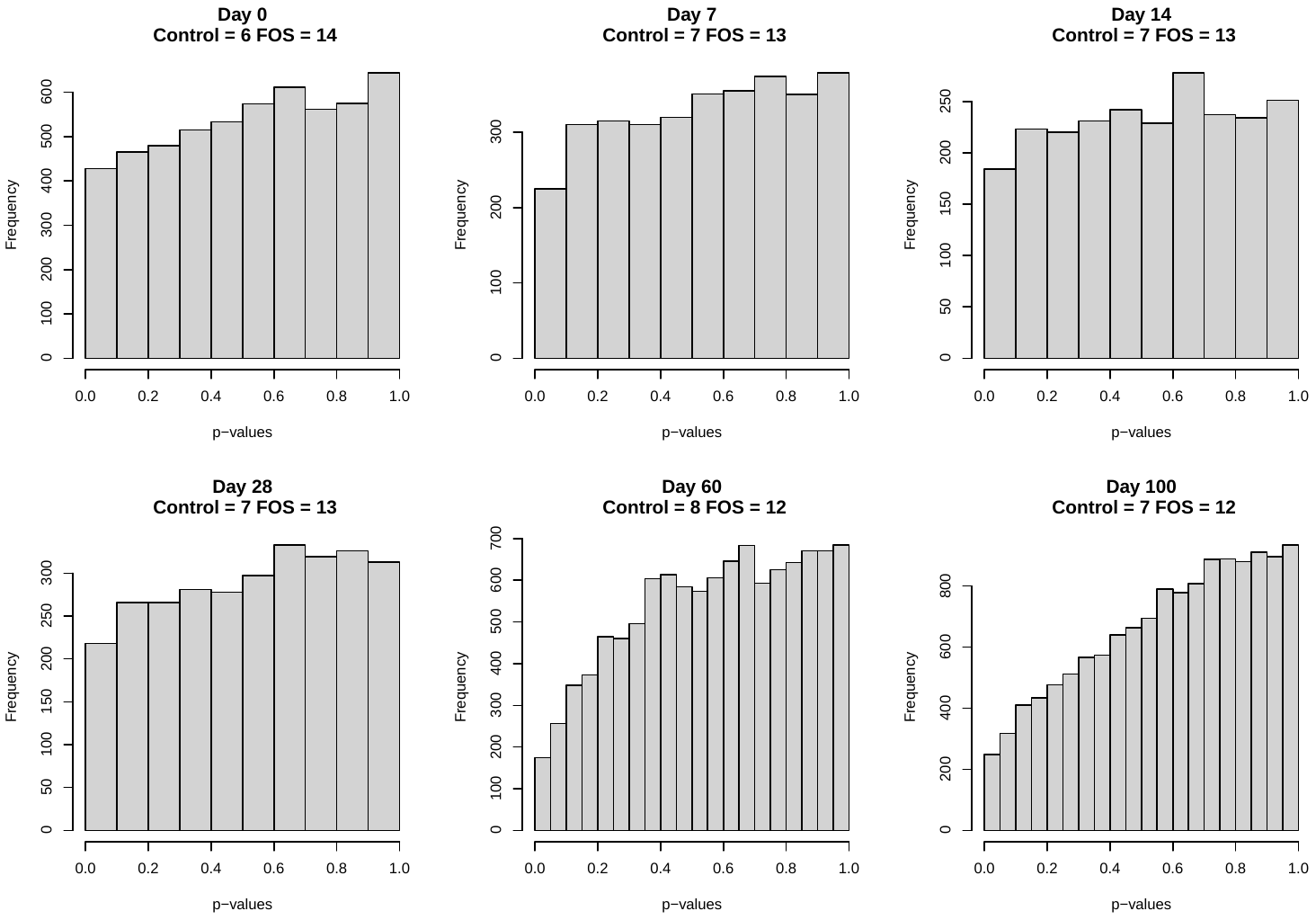

**Supplementary Figure 3. No difference in gut microbiota alterations in response to transplant observed between FOS and control groups**. Histograms showing p-values from Student’s t-test comparing the change in the relative abundance of species at each time point compared to the baseline (day -5) between FOS and control groups. Number of FOS and control samples that were available both at a time point and baseline (day -5) are shown.

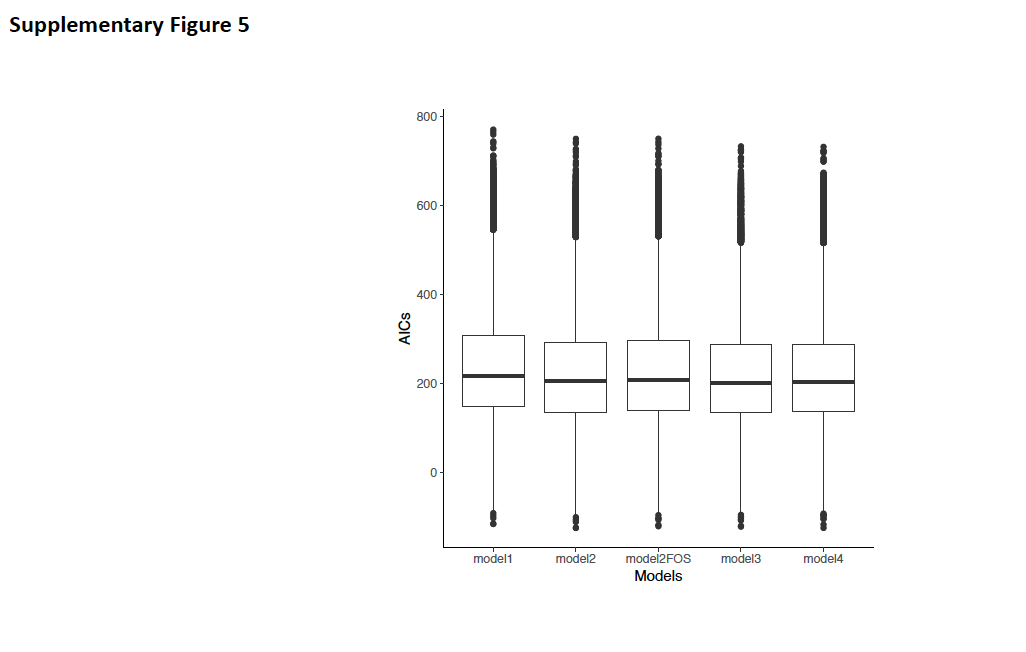

**Supplementary Figure 4.** **AIC analysis indicated that mixed linear model with a second-degree polynomial for time as a fixed effect and patient ID as a random effect was a reasonable fit compared to alternative models to capture changes in the gut microbiota after transplant.** Model 1 (taxa ~ -day, random = ~1 | patient ID) had a significantly higher AIC (mean ± SD: 244.2 ± 142.3) compared to other models (pairwise t-test adjusted p < 0.001). Model2 (taxa ~ -day^2^, random = ~1 | patient ID) (mean AIC ± SD: 230.5 ± 139.9) was comparable to Model2FOS (taxa ~ -day^2^ + FOS, random = ~1 | patient ID) (mean AIC ± SD: 233.5 ± 139.1, adjusted p = 0.09), Model 3 (taxa ~ -day^3^, random = ~1 | patient ID) (mean ± SD: 226.7 ± 137.2, adjusted p = 0.04), and Model 4 (taxa ~ -day^4^, random = ~1 | patient ID) (mean AIC ± SD: 227.5 ± 136.3, adjusted p = 0.09). Model 2 was used for modeling changes in the gut microbiota following transplant since it was simpler and has fewer parameters compared to Model2FOS, Model3, and Model4. (AIC: Akaike information criterion).

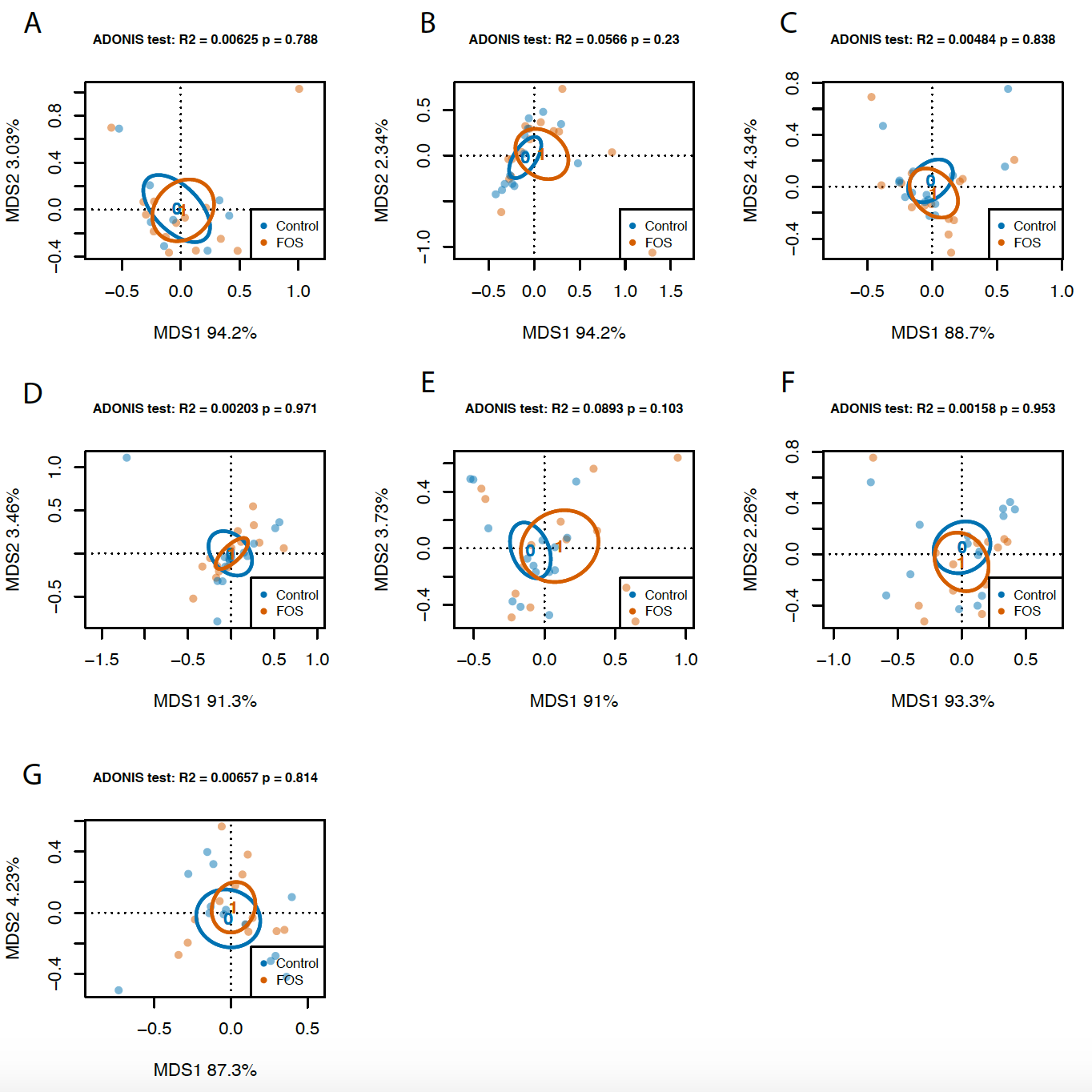

**Supplementary Figure 5.** **Gut microbiota function is not significantly different between FOS and control groups**. Principal coordinate analyses of gut microbiota function were performed using Bray-Curtis distances at each time point (A : day -5, B : day 0, C: day 7, D: day 14, E: day 28, F: day 60, G: day 100). The ADONIS test was used to compare gut microbiota function between FOS and control groups at each time point.

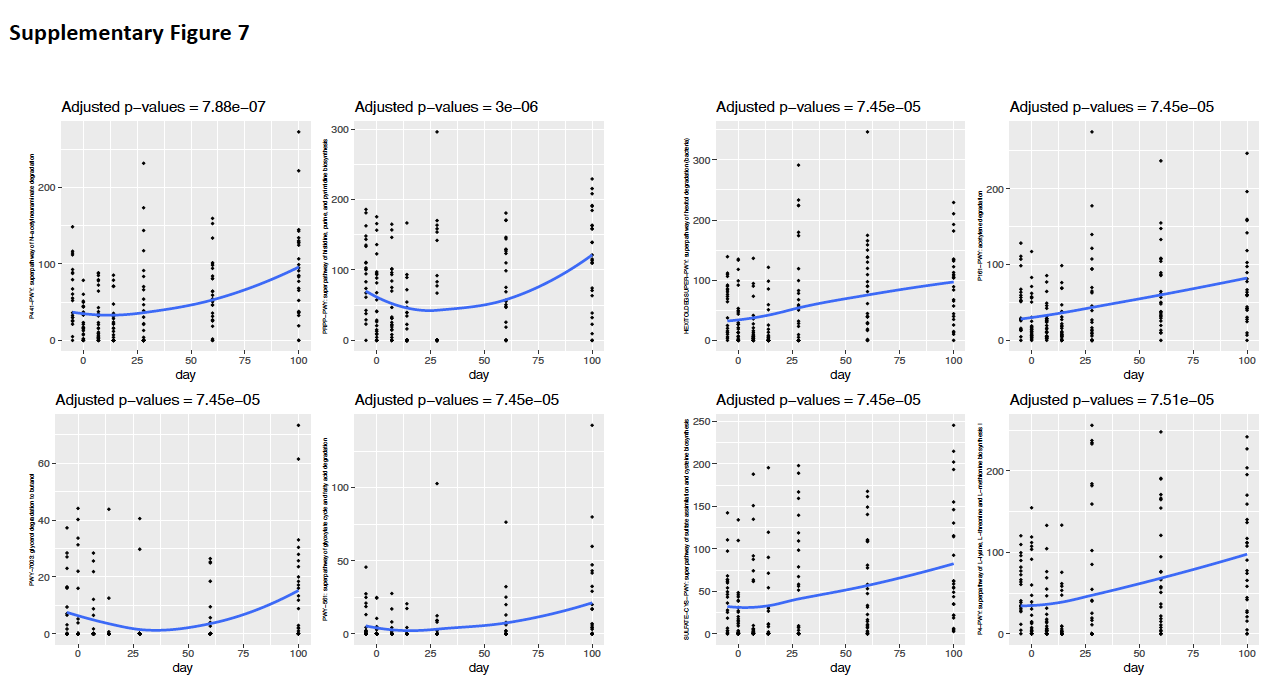

**Supplementary Figure 6.** **Metabolic pathways undergo significant changes following transplantation.** The plots show the relative abundance of the most significant metabolic pathways from mixed linear models with a second-degree polynomial for time as a fixed effect and patient ID as a random effect (taxa ~ -day^2^, random = ~ 1 | patient ID). The y-axis represents normalized count reads (counts per million). The blue line shows the predicted values for the normalized reads at each time point.

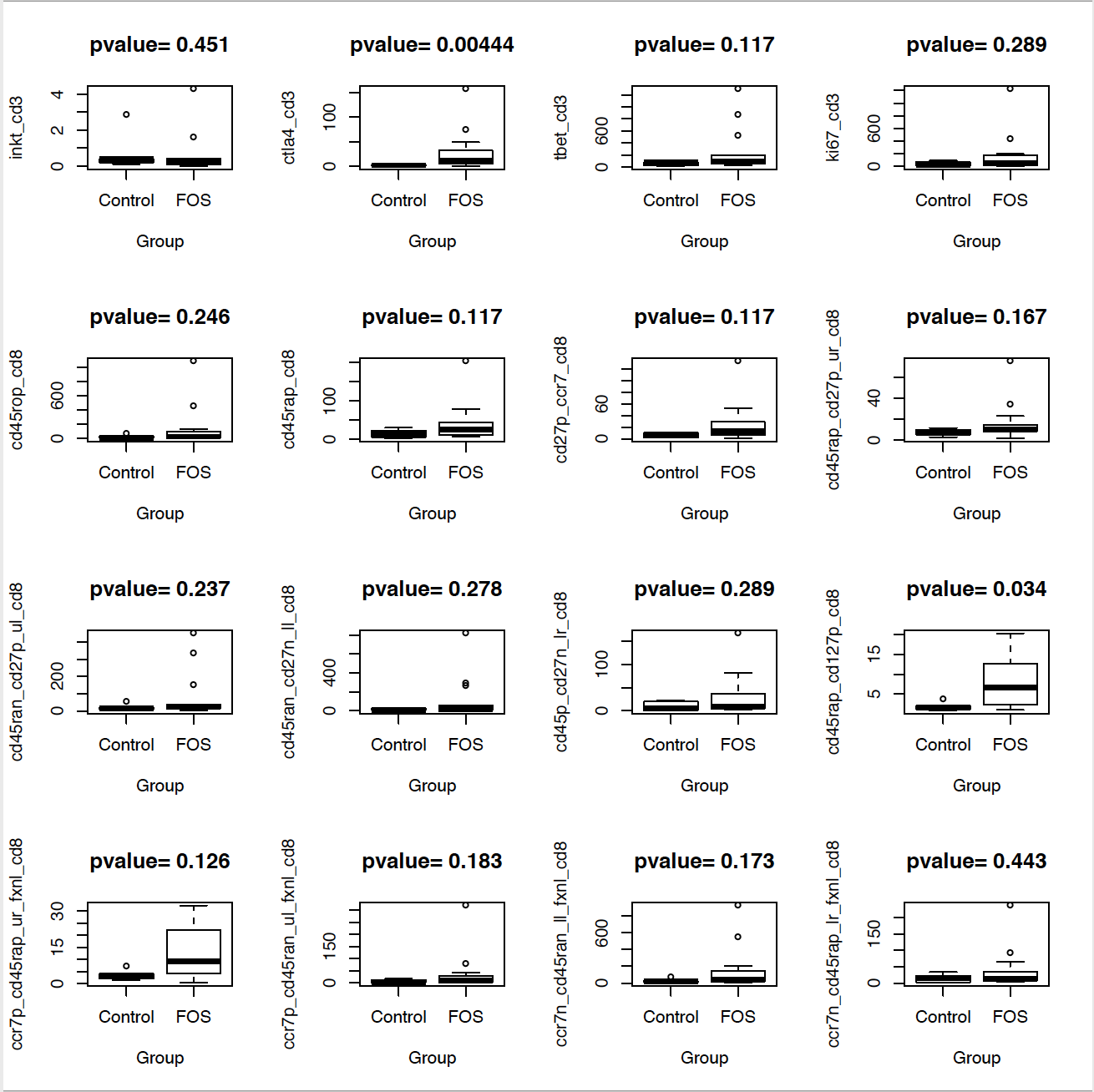

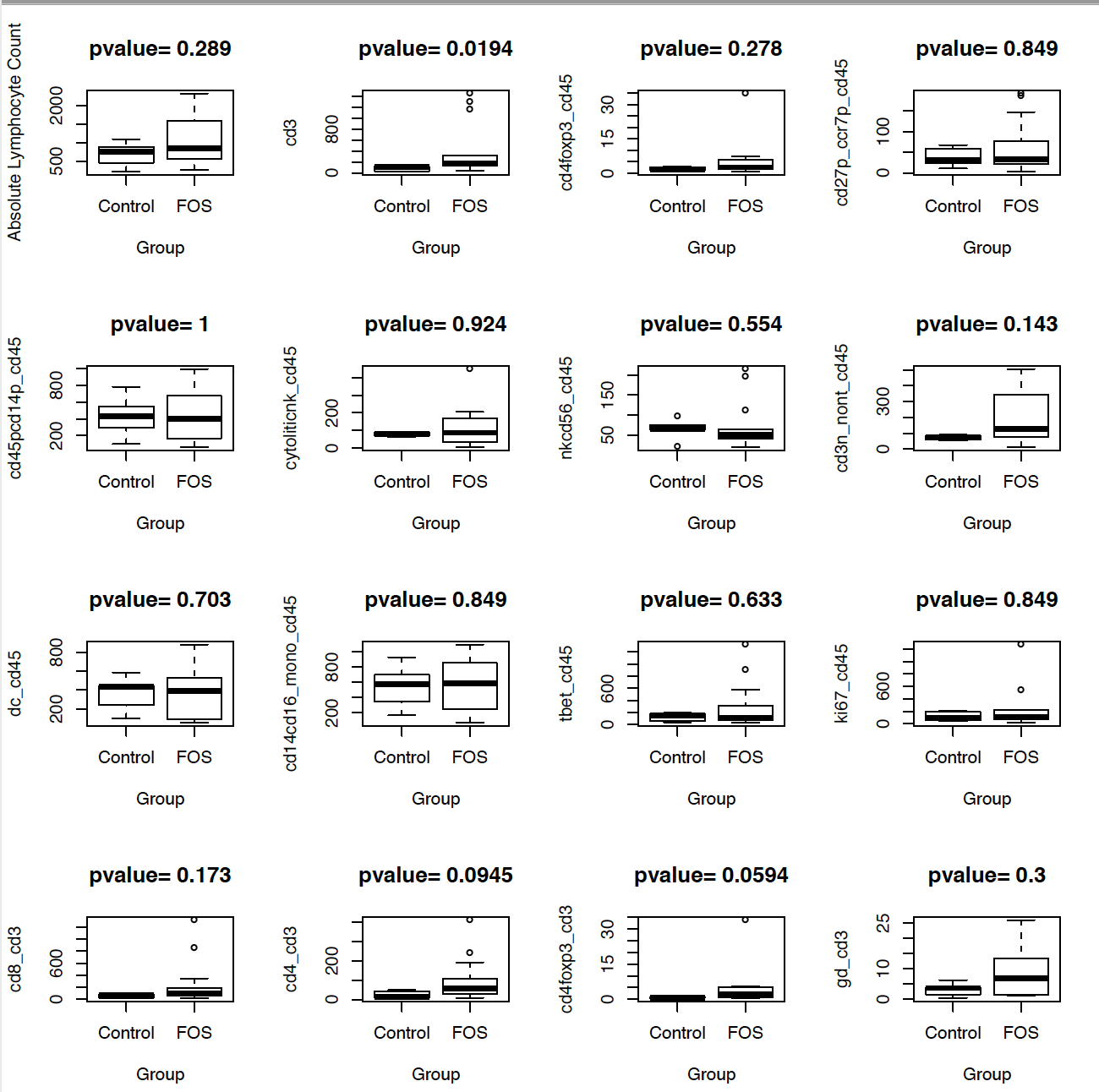

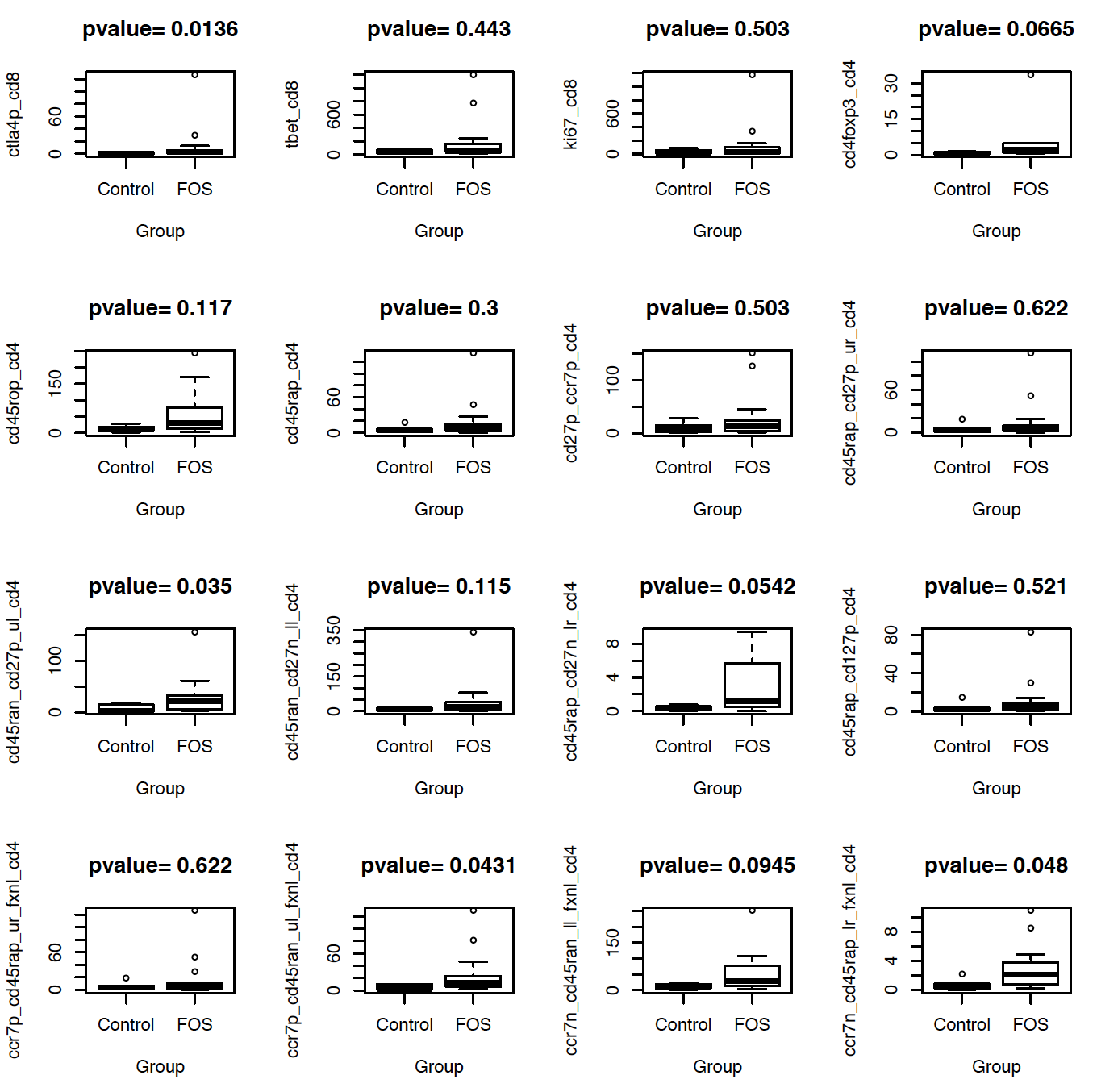

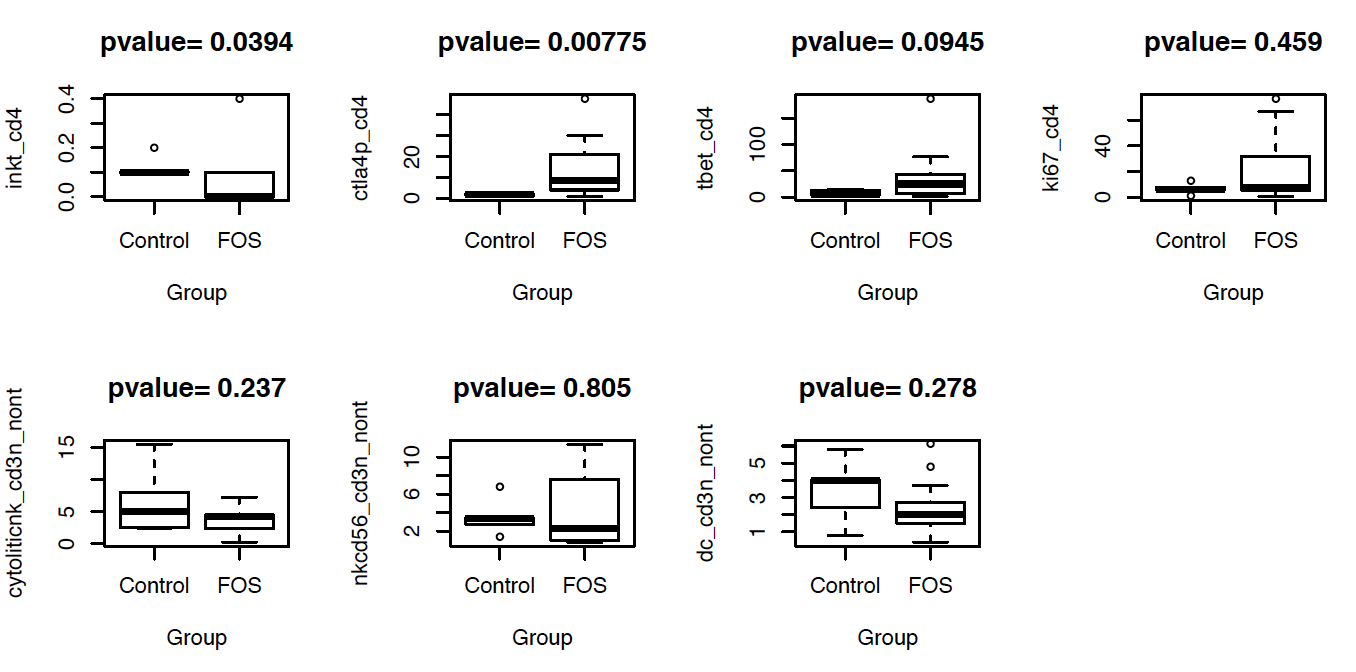

**Supplementary Figure 7. Boxplots comparing CyTOF results between FOS and control groups**. P-values shown are unadjusted; no significant differences were observed in concentrations of T- and other cellular subsets after adjusting using the Benjamini-Hochberg procedure. Highlighted boxplot in red demonstrates the differences in CD4+ FoxP3+ T-regulatory cells between patients receiving FOS compared to controls.
