## Supplementary materials for "A fructo-oligosaccharide prebiotic is well-tolerated in adults undergoing allogeneic hematopoietic stem cell transplantation: a phase I dose-escalation trial"

**Supplementary information**

Statistical design

We will employ the Bayesian optimal interval (BOIN) design to find the maximum tolerable dose under the assumption of a maximum sample size of 15 patients and a target toxicity rate of 35% (65% of patients will be able to tolerate 70% of the doses). The BOIN design is a novel Bayesian dose-finding method that optimizes patient ethics by minimizing the chance of exposing patients to subtherapeutic and overly toxic doses.

The BOIN design yields an average performance comparable to that of the continual reassessment method (CRM) in terms of selecting the MTD, but has a lower risk of assigning patients to sub-therapeutic or overly toxic doses (i.e., better patient ethics). We used the R package version 2.0 software in the development of our study design ([https://CRAN.R-project.org/package=BOIN](https://cran.r-project.org/package=BOIN)).

Given a target rate of 0.35 and a maximum sample size of 15, the BOIN design can be described as follows:

| Number of patients treated | 5 | 10 | 15 |
| --- | --- | --- | --- |
| Escalate if number of DLT* ≤ | 1 | 2 | 4 |
| De-escalate if number of DLT ≥ | 2 | 4 | 6 |
| Eliminate if number of DLT ≥ | 4 | 7 | 9 |

*DLT = dose-limiting toxicity (i.e.  a person who has a DLT does not tolerate that dose)

- Patients in the first cohort are treated at the lowest dose level.
- To assign a dose to the next cohort of patients, we conduct dose escalation/de-escalation according to the rule displayed above, in which “Eliminate” means that we eliminate the current and higher doses from the trial to prevent treating any future patients at these doses because they are overly toxic. When we eliminate a dose, we automatically de-escalate the dose to be assigned to the next lower level. When the lowest dose is eliminated, we stop the trial for safety. In this case, no dose should be selected as the MTD. If none of the actions (i.e., escalation, de-escalation or elimination) is triggered, we treat the new patients at the current dose.
- If the current dose is the lowest dose and the rule indicates dose de-escalation, we will treat the new patients at the lowest dose unless the number of DLTs reaches the elimination boundary, at which point we will terminate the trial for safety. If the current dose is the highest dose and the rule indicates dose escalation, we will treat the new patients at the highest dose.
- Escalate dose if the observed tolerance rate at the current dose >= 0.70 (toxicity <= 0.30 )
- Deescalate dose if the observed tolerance rate at the current dose <=0.60 (toxicity >= 0.40 )

**Appendix**

PRO-CTCAE based questionnaire

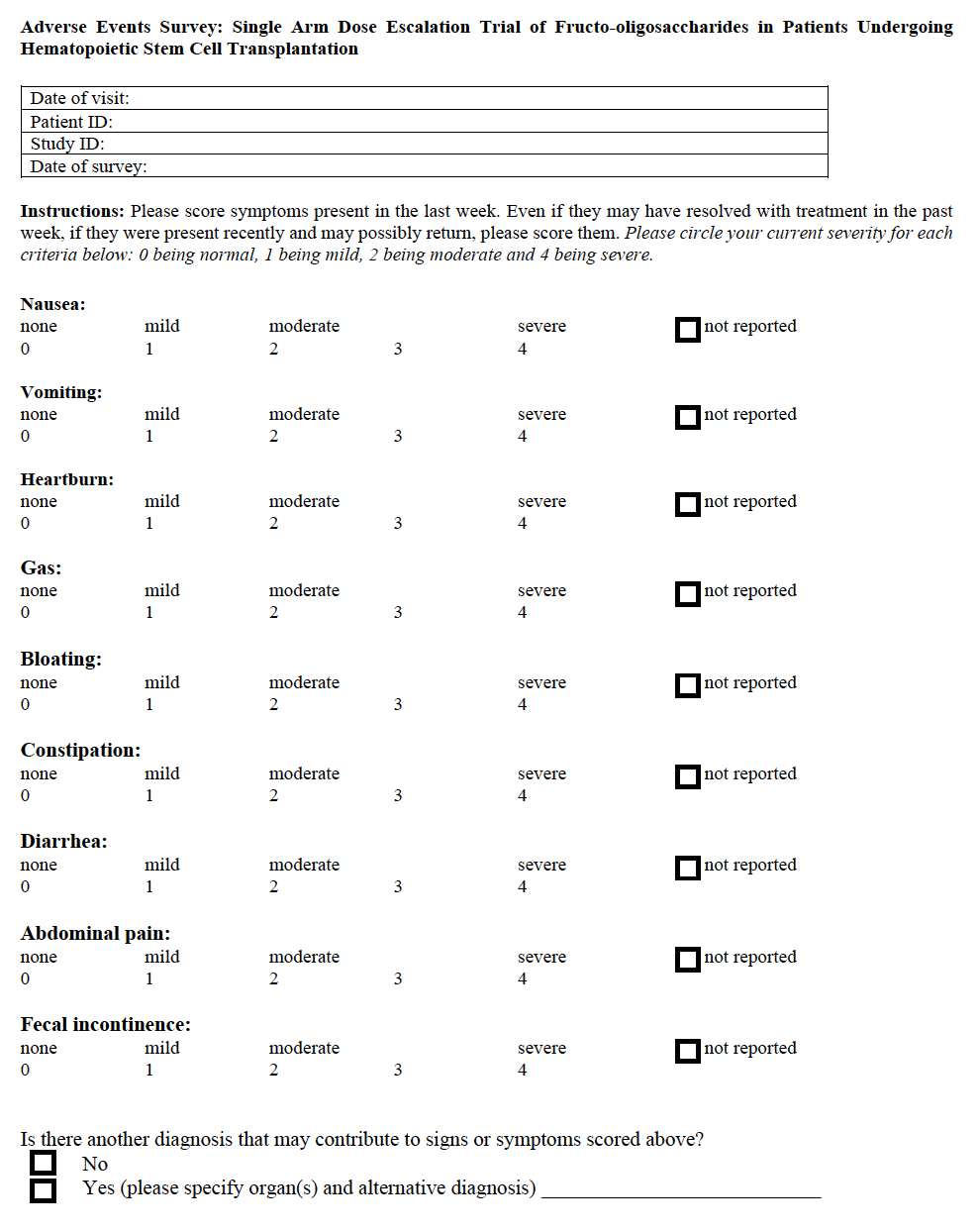
